# Cervical mucosal inflammation expands functional polymorphonuclear myeloid-derived suppressor cells

**DOI:** 10.1101/2024.07.10.24310202

**Authors:** Daan K.J. Pieren, Aleix Benítez-Martínez, Vicente Descalzo, Maider Arando, Patricia Álvarez-López, Jorge N. Garcia-Perez, Núria Massana, Júlia Castellón, Yannick Hoyos-Mallecot, Daniel Alvárez-Sierra, Clara Ramírez-Serra, Nuria Laia Rodriguez, Laura Mañalich-Barrachina, Cristina Centeno-Mediavilla, Josep Castellví, Vicenç Falcó, María J. Buzón, Meritxell Genescà

## Abstract

The mucosal immune system plays a fundamental role in maintaining microbial balance. Microbial imbalance in the female genital tract increases the risk for adverse health outcomes in women and may increase susceptibility to genital tract infections. Among different relevant immune subsets, myeloid-derived suppressor cells (MDSCs) remain understudied in the context of female genital tract conditions. Here we show that frequency of polymorphonuclear (PMN-) MDSCs increased in the cervical mucosa of women with *Chlamydia trachomatis*, bacterial vaginosis, or with a coinfection, but not in women with human papillomavirus. Mucosal PMN-MDSC frequencies correlated with mucosal IL-1β in *C. trachomatis* patients and e*x vivo* exposure of cervical tissue to *C. trachomatis* elevated both PMN-MDSC frequencies and IL-1β secretion. Likewise, exposure of cervical tissue to cervicovaginal lavage fluid from *C. trachomatis* and bacterial vaginosis patients also enhanced PMN-MDSC frequencies. Lastly, cervical MDSCs expressed suppressive mediators and functionally suppressed cytotoxic T-cell responses. Our study identifies IL-1β-stimulated PMN-MDSCs as an immune suppressive mediator in female genital tract infections, potentially contributing to susceptibility to acquiring secondary infections at this site.

## Introduction

Microbial disturbances of the female genital tract (FGT) caused by bacterial vaginosis (BV) and sexually transmitted infections (STIs) have a detrimental impact on women’s health, including genital tract inflammation, infertility, adverse birth outcomes, and cancer (1–3). Furthermore, disorders of the FGT caused by BV (4–6), as well as *Chlamydia trachomatis* (CT) (7), human papillomavirus (HPV) (8, 9), and other pathogenic infections have been shown to enhance the risk of acquiring secondary STIs, such as HIV (10–14). A better understanding of immunological factors in the FGT initiated by primary microbial disturbances is important for development of new vaccination strategies and is vital to identify mechanisms that may contribute to enhanced susceptibility to secondary STIs, thereby aiding overall women’s health.

Myeloid-derived suppressor cells (MDSCs) exert potent suppressive effects on other immune cells and may expand as a consequence of alterations in their cellular micro-environment. MDSCs have widely been reported to expand as a consequence of cancer and infectious diseases and may detrimentally suppress immune responses required for the elimination of tumors and pathogens (15–17). Contrastingly, MDSC-mediated suppression may also be beneficial as it may promote maternal-fetal tolerance during several stages of pregnancy by suppression of fetal-rejecting immune responses (18, 19). Still, not much is known about the presence of MDSCs in the FGT under homeostatic nor pathological conditions.

MDSCs develop under the presence of several cytokines and chemokines, including GM-CSF, VEGF, IL-6, IL-1β, and CFS1 (15), many of which have also been reported to be involved during infection of the FGT with CT and other pathogens (20–22). MDSCs have been classified to mainly consist of two subsets; a subset derived from monocytic myeloid lineages termed monocytic (M-) MDSCs, and a subset derived from granulocytic lineages termed polymorphonuclear (PMN-) MDSCs. These cells contain multiple mechanisms to exert immune suppression, which include expression of the enzyme indoleamine 2,3-dioxygenase (IDO) and the inhibitory ligand Programmed death-ligand 1 (PD-L1) (23). These suppressive factors may limit cytotoxic functions of effector immune cells, such as resident memory T cells, including IFN-γ production and degranulation (24–26), required for effective elimination of infectious pathogens such as CT (27–31). Moreover, pathogen-induced secretion of cytokines in the FGT may attract CD4^+^ T cells towards the cervical mucosa as target cells for HIV, as shown for CT infection (32). Combined with MDSC-mediated suppression of T cell responses in the FGT, these processes may predispose for enhanced HIV susceptibility, negatively affecting women’s health.

Here, we investigated the presence of PMN- and M-MDSCs in the cervical mucosa and blood of women with multiple genital tract conditions. We found higher frequencies of PMN-MDSCs in the cervical mucosa of CT, BV, and coinfected patients associated to local changes that were not reflected in blood. Further, using an *ex vivo* cervical tissue model, we identified IL-1β as a main driver of PMN-MDSC activity in the context of CT infection. Our findings indicate a role for PMN-MDSCs during CT and BV FGT conditions, which contribute to immune suppression at this mucosal site and may thereby limit pathogen clearance and potentially enhance susceptibility to secondary STIs.

## Results

### Study cohort

Cervical cytobrush samples and cervicovaginal lavage fluid were acquired from women suspected for having an STI, in addition to paired PBMC and plasma samples. In total, 74 women were included in this study (**Figure 1A**), from which part did not have a genital infection (Healthy donors, HD, n=18), or had a laboratory confirmed human papillomavirus infection (HPV, n=19) or *Chlamydia trachomatis* infection (CT, n=14), or had bacterial vaginosis determined by Ison-Hay criteria (BV, n=12), or a coinfection of CT, BV, *Neisseria gonorrhoeae*, *Mycoplasma genitalium*, and/or *Candida albicans* (Coinf, n=11). Patient group characteristics including age, phase of the menstrual cycle and other clinical parameters are presented in **Supplemental Table 1**. Of note, women assigned to the CT and Coinf groups were in general younger than women in the other groups, while the percentage of patients using hormonal contraceptives did not differ between groups (**Supplemental Table 1**). Pathogens identified in each individual patient of the coinfection group are presented in **Supplemental Table 2**.

**Figure 1.**
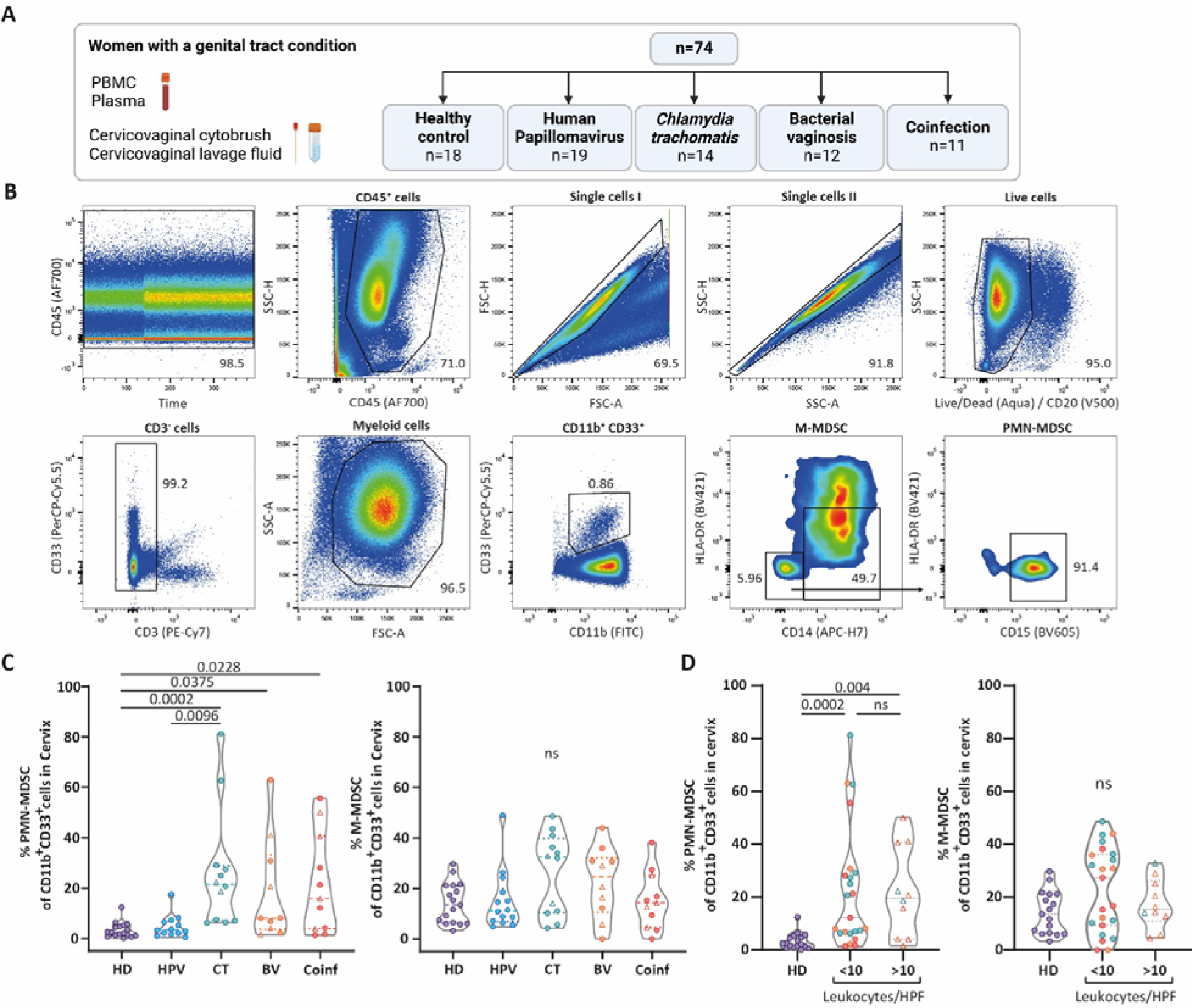
PMN-MDSC frequencies are increased in the cervical mucosa of Chlamydial, bacterial vaginosis, and coinfected patients. (A) Schematic overview of study participants and sample collection. (B) Representative flow- cytometry plots showing the gating strategy of M-MDSCs (CD33^+^CD11b^+^HLA-DR^dim^CD14^+^ myeloid cells) and PMN-MDSCs (CD33^+^CD11b^+^HLA-DR^-^CD14^-^CD15^+^ myeloid cells) in cervical samples. (C,D) Comparison of PMN-MDSC and M-MDSC frequencies amongst CD11b^+^CD33^+^ myeloid cells found in cervical samples of (C) each study group (HD n=18; HPV n=13; CT n=12; BV n=10; Coinf n=11) and in (D) groups according to leukocyte/High Power Field (HPF) count (HD n=18; <10 leukocytes/HPF n=23; >10 leukocytes/HPF n=10). Data are shown as violin plots with median and quartiles (C,D). Patients with leukocyte/HPF counts of >10 are indicated with triangles. Patient characteristics are shown in Supplementary Table 1. Statistical significance was determined by Kruskal-Wallis test with Dunn’s post hoc test (two-sided). Exact *P*-values are shown; ns not significant.

### PMN-MDSC frequencies are increased in the cervical mucosa of women with C. trachomatis, bacterial vaginosis, and coinfections

To investigate the presence of MDSCs during FGT homeostatic and microbially altered conditions, we assessed the frequency of M-MDSCs and PMN-MDSCs in cervical and PBMC samples of recruited participants by flow cytometry. Whereas myeloid cell population is highly heterogeneous and current phenotypical markers do not allow for stringent identification of *bona-fide* MDSC subsets, we applied widely used phenotypical markers to identify MDSC subsets in these samples (23). We characterized M-MDSCs as CD33^+^CD11b^+^HLA-DR^dim^CD14^+^ cells, and PMN-MDSCs as CD33^+^CD11b^+^HLA-DR^-^ CD14^-^CD15^+^ cells (23) in cervical (**Figure 1B**) and PBMC (**Figure 2A**) samples, after gating for single viable myeloid cells and exclusion of CD3^+^ and CD20^+^ cells. Of note, since we did not eliminate neutrophils by gradient centrifugation in cervical samples in order to avoid cell lost, we were not able to exclude potential contamination of neutrophils in these samples. Still, suppressive neutrophils have been described as CD33^-^ cells (33), indicating exclusion of these cells based on our gating strategy.

**Figure 2.**
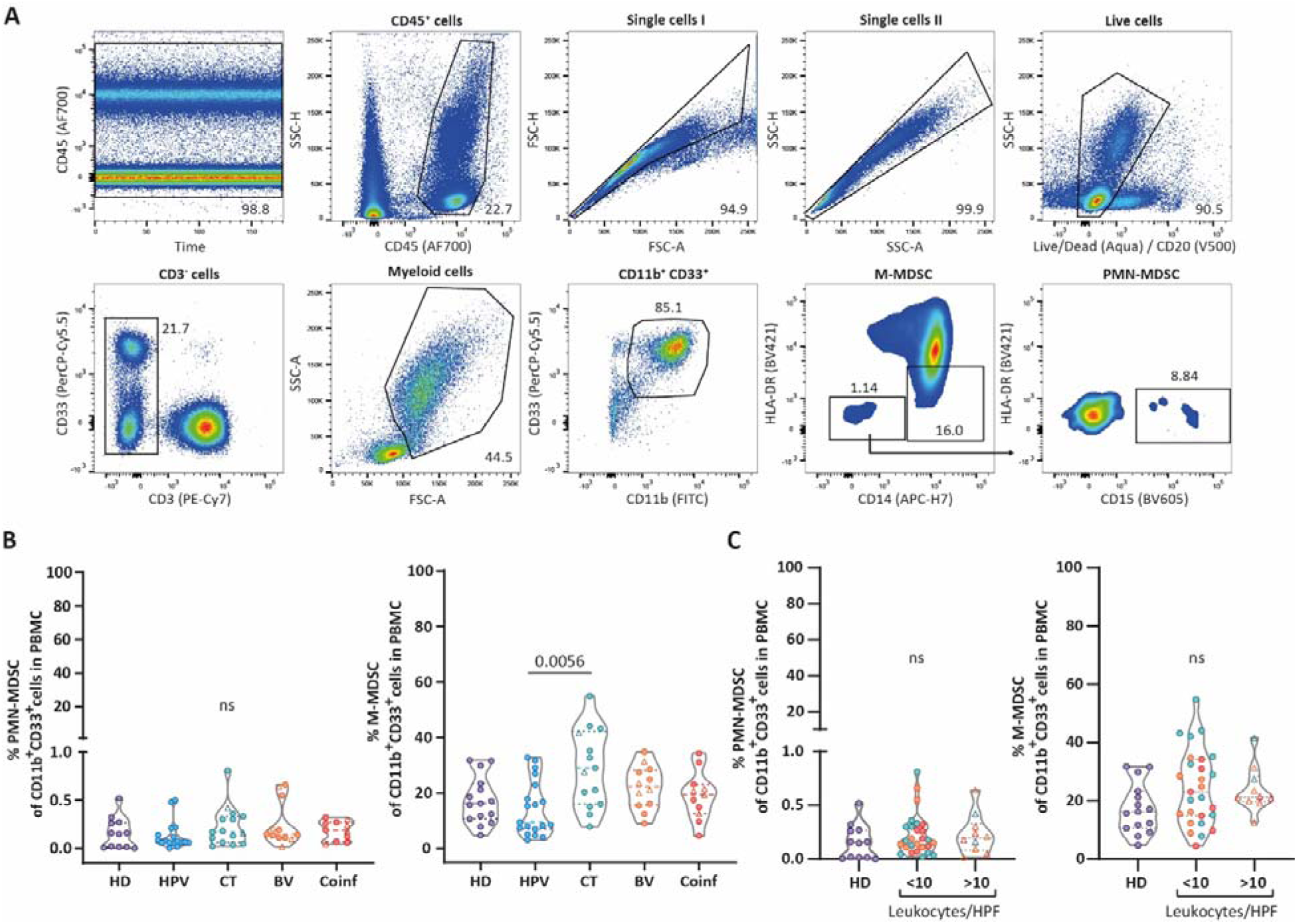
PMN-MDSC frequencies do not change in PBMCs of patients with inflammatory FGT conditions. (A) Representative flow-cytometry plots showing the gating strategy of M-MDSCs (CD33^+^CD11b^+^HLA-DR^dim^CD14^+^ myeloid cells) and PMN-MDSCs (CD33^+^CD11b^+^HLA-DR^-^CD14^-^ CD15^+^ myeloid cells) in PBMC samples. (B,C) Comparison of PMN-MDSC and M-MDSC frequencies amongst CD11b^+^CD33^+^ myeloid cells found in PBMC samples of (B) each study group (HD n=15; HPV n=20; CT n=14; BV n=12; Coinf n=11) and in (D) group according to leukocyte/HPF count (HD n=15; <10 leukocytes/HPF n=27; >10 leukocytes/HPF n=10). Data are shown as violin plots with median and quartiles (B). Patients with leukocyte/HPF counts of >10 are indicated with triangles. Patient characteristics are shown in Supplementary Table 1. Statistical significance was determined by Kruskal-Wallis test with Dunn’s post hoc test (two-sided). Exact *P*-values are shown; ns not significant.

In the cervical mucosa, we found that the frequency of PMN-MDSCs was increased in BV, CT, and Coinf patients compared to HD, while levels in HPV patients remained similar to the HD group (**Figure 1C**). In contrast, M-MDSC levels did not change among the different groups (**Figure 1C**). Several participants across the CT/BV/Coinf groups showed >10 leukocytes per high power field (HPF), indicative for increased inflammation (**Supplemental Table 1**), yet differences observed in cervical PMN-MDSC levels remained regardless of the leukocyte/HPF count (**Figure 1D**). PMN-MDSC and M-MDSC frequencies in PBMC samples were similar among the groups, aside of a significant increase of M-MDSC levels in CT patients compared to HPV patients (**Figure 2B**), and did not change according to leukocyte/HPF count (**Figure 2C**). Frequencies of M-MDSCs were comparable between PBMC and cervical compartments, whereas PMN-MDSC levels were higher in the cervical mucosa (**Supplemental Figure 1A**).

Age showed a negative association with cervical PMN- but not M-MDSC frequencies amongst all patients (**Supplemental Figure 1B**), which was likely an effect of the younger age of the CT and Coinf patients included. To disentangle the effect of age on the observed differences, we performed a post-hoc analysis to determine the frequency of cervical PMN-MDSC with respect to age. When all samples were separated into patients who were above or below the median age (30 years old) no differences were observed (**Supplemental Figure 1C**). In addition, pooling samples from groups with increased cervical PMN-MDSC frequencies, namely CT/BV/Coinf groups, and compared them to pooled HD/HPV groups, differences were maintained regardless of being younger or older than 30 years old (**Supplemental Figure 1C**). Further, menstrual cycle stage, comparing progesterone high with low dominance (week 1/2 versus 3/4), did not appear to influence PMN- and M-MDSC levels, yet higher levels of PMN-MDSCs in the pooled CT/BV/Coinf versus HD/HPV groups were maintained regardless of the menstrual cycle phase (**Supplemental Figure 2A**). Last, use of hormonal contraceptives showed higher levels of cervical PMN-MDSCs, yet (a trend towards) increased PMN-MDSC levels were maintained within pooled CT/BV/Coinf participants compared to HD/HPV groups in the absence or presence of hormonal contraceptives (**Supplemental Figure 2B**).

Additionally, we addressed the presence of several other cell subsets relevant for pathogen clearance in these same samples (**Supplemental Figure 3, 4**), since these populations could similarly be modulated by the microenvironment and potentially interact with MDSC subsets. The CT group exhibited more changes compared to HD than any other group: in the cervical compartment, the frequency of neutrophils (CD33^-^ CD11b^+^ myeloid cells) and the expression of CD69 in CD56^dim^CD16^+^ NK cells were higher (**Supplemental Figure 3B**); while in PBMCs, expression of CD69 was elevated in CD56^hi^CD16^-^ NK cells and lower frequencies of HLA-DR^+^ expression were detected in both NK cell subsets (**Supplemental Figure 4B**). Similarly, BV and Coinf groups showed increased expression of CD69 in CD56^hi^CD16^-^ NK cells from PBMCs only, while lower frequencies of HLA-DR^+^ expression were detected in this same subset only in the BV group (**Supplemental Figure 4B**). Last, the HPV group showed enhanced T-cell expression of HLA-DR in the cervical samples compared to HD (**Supplemental Figure 3B**), while no changes in other subsets from PMBCs were observed for these women (**Supplemental Figure 4B**). Together, we identify increased frequencies of PMN-MDSC associated with BV, CT, and coinfection that can only be detected at the local mucosa.

### Cytokine profiles in CVL fluid of women with genital tract conditions

Multiple soluble factors have been associated to MDSC development, expansion, and suppressive activity, including growth factors (CSF1, G-CSF, GM-CSF, and VEGF-A), inflammatory cytokines (IL-1β, IL-6, and TNF-α) and an anti-inflammatory cytokine (TGF-β1) (15). Thus, we next measured the levels of these factors in plasma and cervicovaginal lavage (CVL) fluid from our patients. BV patients showed higher levels of CSF1, GM-CSF, VEGF-A, IL-1β, and TGF-β1 compared to HD (**Figure 3A**), whereas CT patients only showed higher levels of CSF1 compared to HD. Coinf patients showed increased levels of CSF1, but also GM-CSF, and IL-1β, while HPV patients did not show any differences compared to HD (**Figure 3A**). High dispersion levels observed for some of these cytokines did not appear to be influenced by the number (<10 or >10) of leukocytes/HPF detected (**Supplemental Figure 5A**). In addition, menstrual cycle stage or the use of hormonal contraceptives did not alter CVL fluid cytokine levels (**Supplemental Figure 2A, 2B**). In contrast to CVL fluid, cytokine levels in plasma samples did not show major differences between HD and the other groups, aside from an increased level of TGF-β1 in the Coinf group (**Supplemental Figure 5B**).

**Figure 3.**
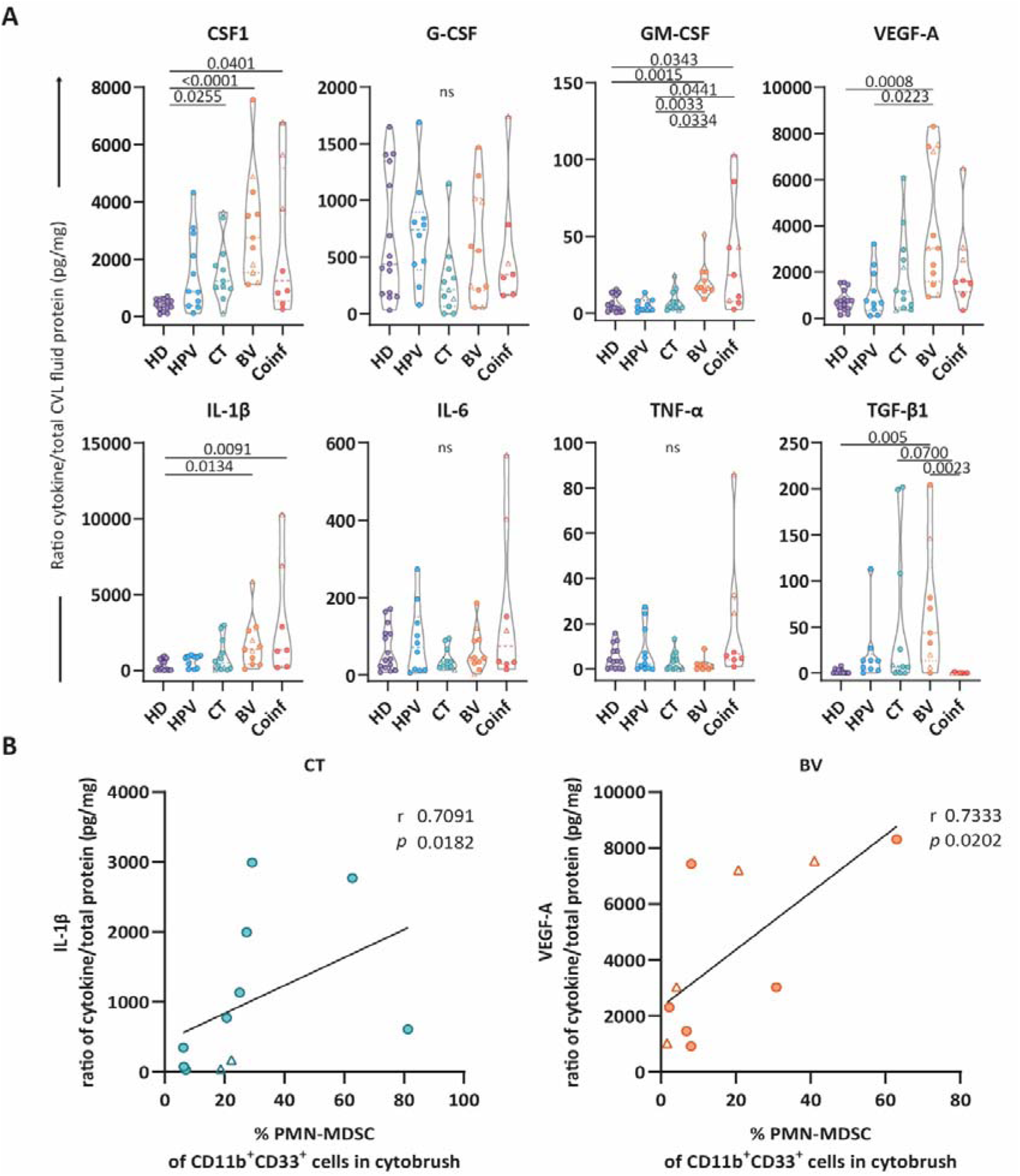
Cytokine and growth factors in cervical lavage fluid. (A) Comparison of levels of eight different growth factors and cytokines found in CVL fluid for each group of women included in this study depicted as the ratio of each cytokine per total protein present in the cervical lavage sample (HD n=14; HPV n=12; CT n=11; BV n=11; Coinf n=9). (B) Graphs show the relationship between the level of CVL fluid IL-1β and the frequency of PMN-MDSCs in the cervical samples from CT patients (n=11) and between the level of CVL fluid VEGF-A and the frequency of PMN-MDSCs in the cervical samples from BV patients (n=10). Data are shown as violin plots with median and quartiles (A) and correlations (*r* and *P* values) assessed by Spearman test (two-sided) (B). Patients with leukocyte/HPF counts of >10 are indicated with triangles. Patient characteristics are shown in Supplementary Table 1. Statistical significance was determined by Kruskal-Wallis test with Dunn’s post hoc test (two-sided) (A). Exact *P*-values are shown, ns not significant.

Next, we assessed whether levels of specific cytokines in the CVL fluid associated with higher frequencies of PMN-MDSCs detected in the cervical mucosa of BV, CT, and Coinf patients as observed in **Figure 1C**. Amongst all patients included in our study, only the level of CSF1 present in CVL fluid positively correlated with the frequency of PMN-MDSCs in the cervical mucosa (*r* 0.45, *p*<0.001) (**Supplemental Table 3**). However, when analyzing within each group of patients, only HPV reflected this finding (*r* 0.74, *p*=0.04). Aside from negative correlations within the HD group, likely caused by low levels of cytokines and low PMN-MDSC frequencies (**Supplemental Table 3**), we observed strong positive correlations between the level of IL-1β found in the CVL of CT patients and the frequency of PMN-MDSCs, while for BV patients the level of VEGF-A positively correlated with the frequency of PMN-MDSCs (**Figure 3B**). Together these data suggest that IL-1β in CT patients and VEGF-A in BV patients could, in part, contribute to the accumulation of PMN-MDSCs found in the cervical mucosa.

### Ex vivo exposure of cervical tissue to C. trachomatis elementary bodies expands the PMN-MDSC subset and increases levels of IL-1***β***

As we detected higher frequencies of PMN-MDSCs in the cervical mucosa of CT patients, we next explored whether *ex vivo* exposure of cervical tissue to infectious elementary bodies (EBs) of *C. trachomatis* could expand PMN-MDSCs. Exposure of cervical tissue to EBs induced higher frequencies of PMN-MDSCs compared to the control (**Figure 4A, 4B**). Exposure to GM-CSF and IL- 6, previously reported as MDSC developmental factors (15) and chosen based on preliminary cytokine stimulation experiments, also induced a trend towards higher levels of PMN-MDSCs (**Figure 4A, 4C**).

**Figure 4.**
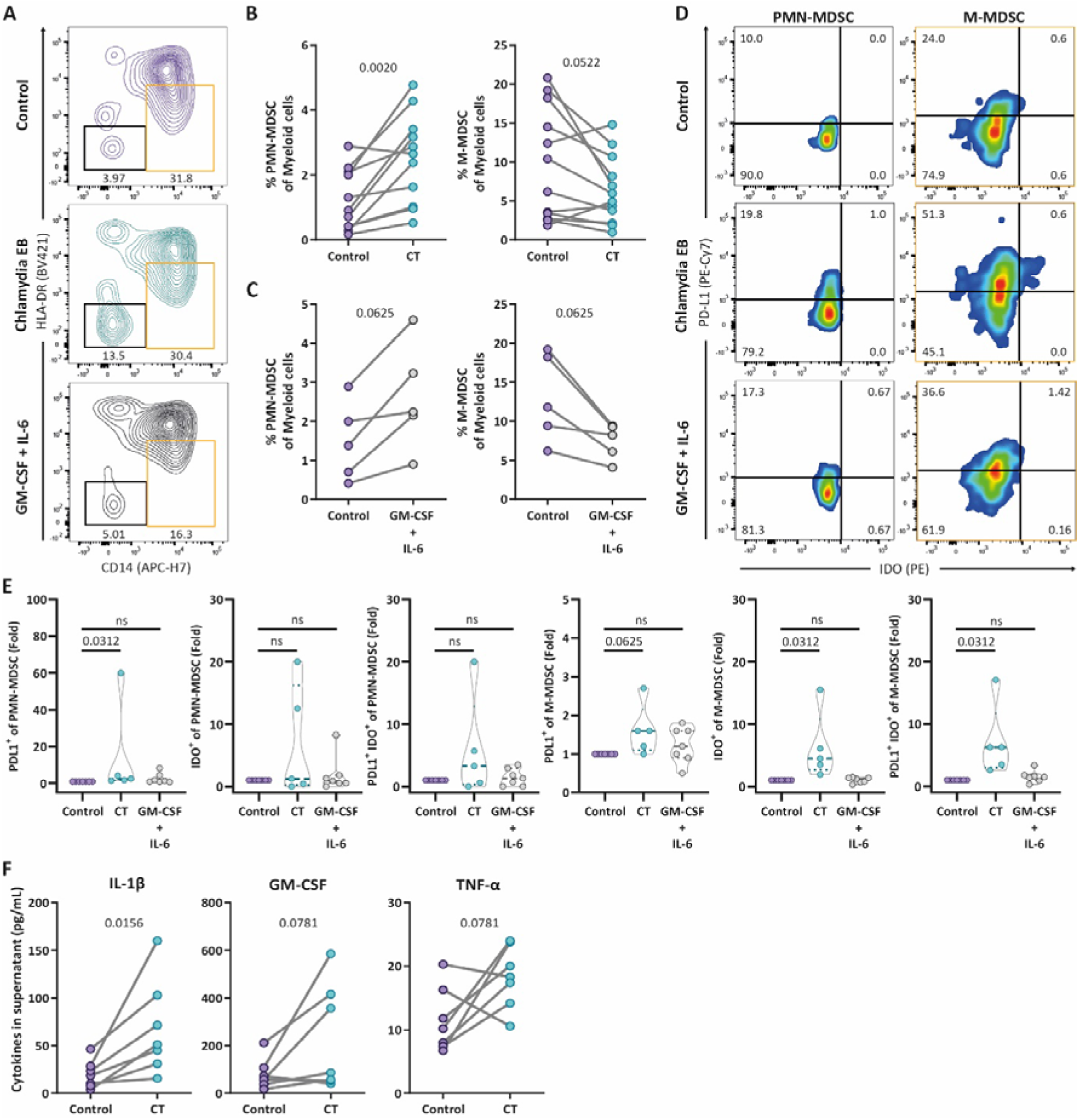
Ex vivo exposure of cervical tissue to C. trachomatis elementary bodies expands the PMN-MDSC subset and increases levels of IL-1β. (A) Representative flow-cytometry plots showing expression of HLA-DR and CD14 to identify PMN- MDSCs and M-MDSCs after exposure of cervical tissue to *C. trachomatis* elementary bodies (EBs), GM-CSF+IL-6, or control for 48 hours. PMN-MDSCs were confirmed to be CD15^+^ after gating for HLA-DR^-^ CD14^-^ cells shown in the plot. (B, C) Comparison of the frequency of PMN-MDSCs and M- MDSCs amongst myeloid cells in cervical tissue exposed to CT EBs (n=11) (B) or GM-CSF+IL-6 (n=5) (C). (D) Representative flow-cytometry plots showing expression of PD-L1 and IDO in PMN- MDSCs and M-MDSCs residing in cervical tissue after exposure to CT EBs for 48 hours compared to the control. (E) Fold change induction of PD-L1^+^, IDO^+^, and PD-L1^+^IDO^+^ PMN-MDSCs and M- MDSCs residing in cervical tissue after exposure to CT EBs or GM-CSF+IL-6 for 48 hours (n=5-7) compared to the control. (F) Comparison of IL-1β, GM-CSF, and TNF-α levels measured in culture supernatant of cervical tissue control *versus* tissue exposed to CT EBs (n=7). Data are shown as paired samples (B, C, F) or violin plots with median and quartiles (E). Statistical significance was determined by Wilcoxon test (two-sided) (B-C and F), or Wilcoxon test (one-sided) in fold change graphs (E). Exact *P*-values are shown, ns not significant.

In contrast, the frequency of M-MDSCs showed a trend towards lower levels in cervical tissue as a consequence of exposure to either EBs or GM-CSF with IL-6 (**Figure 4A-C**). Furthermore, exposure of cervical tissue to CT EBs altered the suppressive phenotype of tissue-derived MDSCs, which we assessed by detection of PD-L1 and IDO expression (23) and verified by FMO controls (**Supplemental Figure 6**). CT EBs induced higher levels of PD-L1^+^ PMN-MDSCs and a trend for M-MDSCs (**Figure 4D, 4E**), while M-MDSCs but not PMN-MDSCs also showed higher levels of IDO^+^ and IDO^+^PD-L1^+^ cells (**Figure 4D, 4E**). In contrast, GM-CSF with IL-6 did not induce changes to the suppressive phenotype of these cells. These data indicate that CT EBs may expand PMN-MDSCs, while also potentially enhancing the suppressive capacity of PMN- and M-MDSCs.

We then measured the level of cytokine concentrations in the culture supernatant to address whether secreted signals were similar to the cervical milieu detected in CT patients, thus potentially explaining expansion of PMN-MDSCs in EB-exposed cervical tissue. The level of IL-1β was significantly higher upon exposure to EBs compared to the control (**Figure 4F**), in agreement with the correlation found in the CVL fluid of CT patients (**Figure 3B**), suggesting a potential role for this cytokine. Additionally, levels of GM-CSF and TNF-α showed a trend towards higher levels upon exposure to EBs compared to the control (**Figure 4F**), whereas levels of CSF1, G-CSF, VEGF-A, IL-6, and TGF-β1 were not modified (**Supplemental Figure 7**). Together, our data indicate that CT induces expansion of PMN-MDSCs, expression of phenotypical markers related to immune suppression on both PMN- and M-MDSCs, and increased levels of IL-1β.

### CVL fluid of CT and BV patients and IL-1***β*** exposure recapitulate PMN-MDSC expansion in cervical tissue

To further investigate the link between the FGT cytokine micro-environment and MDSC expansion and/or activity, we addressed whether CVL fluid from CT or BV patients or IL-1β were capable of inducing expansion of PMN-MDSCs in cervical tissue. To this end, cervical tissue blocks were exposed to medium supplemented with CVL fluid from CT or BV patients, IL-1β, or HD CVL as a control for 24 hours. CVL fluid of CT and BV patients induced higher levels of PMN-MDSCs in cervical tissue compared to tissue exposed to HD CVL fluid (**Figure 5A, 5B**). In contrast, tissue M-MDSC levels did not change in response to CT CVL fluid, while M-MDSC levels decreased when exposed to BV CVL fluid (**Figure 5A, 5B**). Similar to CT CVL fluid, direct exposure of cervical tissue to IL-1β resulted in a trend towards increased PMN-MDSC levels compared to the HD control (*P* 0.0547), but not M-MDSC levels (**Figure 5B**).

**Figure 5.**
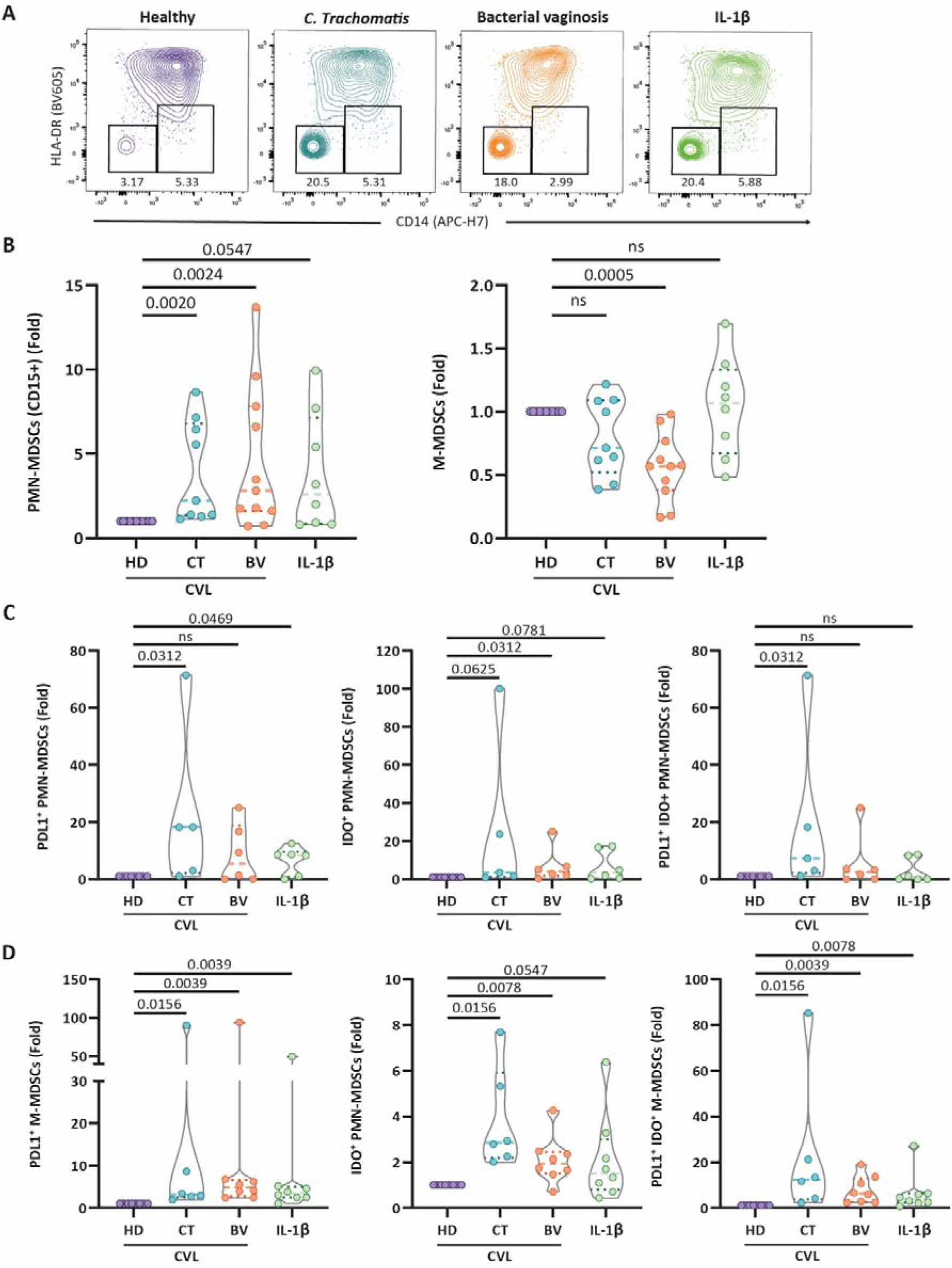
CVL fluid of CT and BV patients and IL-1β exposure recapitulate PMN-MDSC expansion and expression of PD-L1/IDO in cervical tissue. (A) Representative flow-cytometry plots showing expression of HLA-DR and CD14 to identify PMN-MDSCs and M-MDSCs after overnight exposure of cervical tissue to CVL fluid derived from HD, CT, or BV patients, or IL-1β. PMN-MDSCs were confirmed to be CD15^+^ after gating for HLA-DR^-^ CD14^-^ cells shown in the plot. (B) Fold change induction of PMN-MDSCs and M-MDSCs amongst CD11b^+^CD33^+^ myeloid cells in cervical tissue exposed to CVL fluid CT (n=9), or BV (n=11) patients, or IL-1β (n=8) for 24 hours compared to HD CVL fluid. Fold change induction of PD-L1^+^, IDO^+^, and PD-L1^+^IDO^+^ amongst PMN-MDSCs (C) and M-MDSCs (D) in cervical tissue exposed to CVL fluid of CT (n=5-6), or BV patients (n=6-8), or IL-1β (n=6-8) for 24 hours compared to HD CVL fluid (n=6-8). Data are shown as violin plots with median and quartiles (B-D). Statistical significance was determined by Wilcoxon test (one-sided) in fold change graphs (B-D). Exact *P*-values are shown; ns not significant.

Furthermore, CT CVL fluid induced higher levels of PD-L1^+^and PDL1^+^IDO^+^ PMN-MDSCs, and a trend towards higher levels of IDO^+^ PMN-MDSCs (*P* 0.0625) (**Figure 5C**). Exposure to BV CVL fluid resulted in higher levels of IDO^+^ PMN-MDSCs only, while IL-1β induced higher levels of PD- L1^+^ and a trend towards higher IDO^+^ PMN-MDSC levels (*P* 0.0781) (**Figure 5C**). In addition, M-MDSCs showed higher levels of PD-L1^+^, IDO^+^, and PD-L1^+^IDO^+^ cells in response to CT and BV- derived CVL fluid, as well as IL-1β (**Figure 5D**). Together, these findings indicate that CVL fluid of CT and BV patients may contain a specific cytokine micro-environment that results in increased frequencies of PMN-MDSCs with enhanced suppressive potential, which is partially driven by presence of IL-1β.

### Cervical MDSCs suppress T-cell activation and cytotoxic potential

MDSCs isolated from peripheral blood have previously been shown to suppress T-cell cytotoxic responses and proliferation (24, 25, 34). To functionally confirm the suppressive activity of MDSCs residing in cervical tissue we determined the capacity of MDSCs to suppress expression of IFN-γ, granzyme B and degranulation marker CD107a by T cells, after stimulation with anti-CD3/anti-CD28 beads overnight with or without the presence of MDSCs (**Figure 6A**). While these experiments were highly limited by cell yield, when possible, we included cervical neutrophils as a control. Addition of MDSCs to the T-cell cultures resulted in suppression of CD8^+^ and CD4^+^ T cells degranulating (CD107a^hi^) in combination with expression of CD69 or granzyme B (**Figure 6B**). Additionally, MDSCs suppressed the frequency of CD107a^+^ CD8^+^ T cells while in some samples IFN-γ^+^ CD4^+^ T cells (in three out of five) and granzyme B^+^ CD4^+^ T cell frequencies (in three out of six) were also suppressed. In comparison, coculture with cervical neutrophils did not consistently suppress T cell function and activation (**Figure 6B)**.

**Figure 6.**
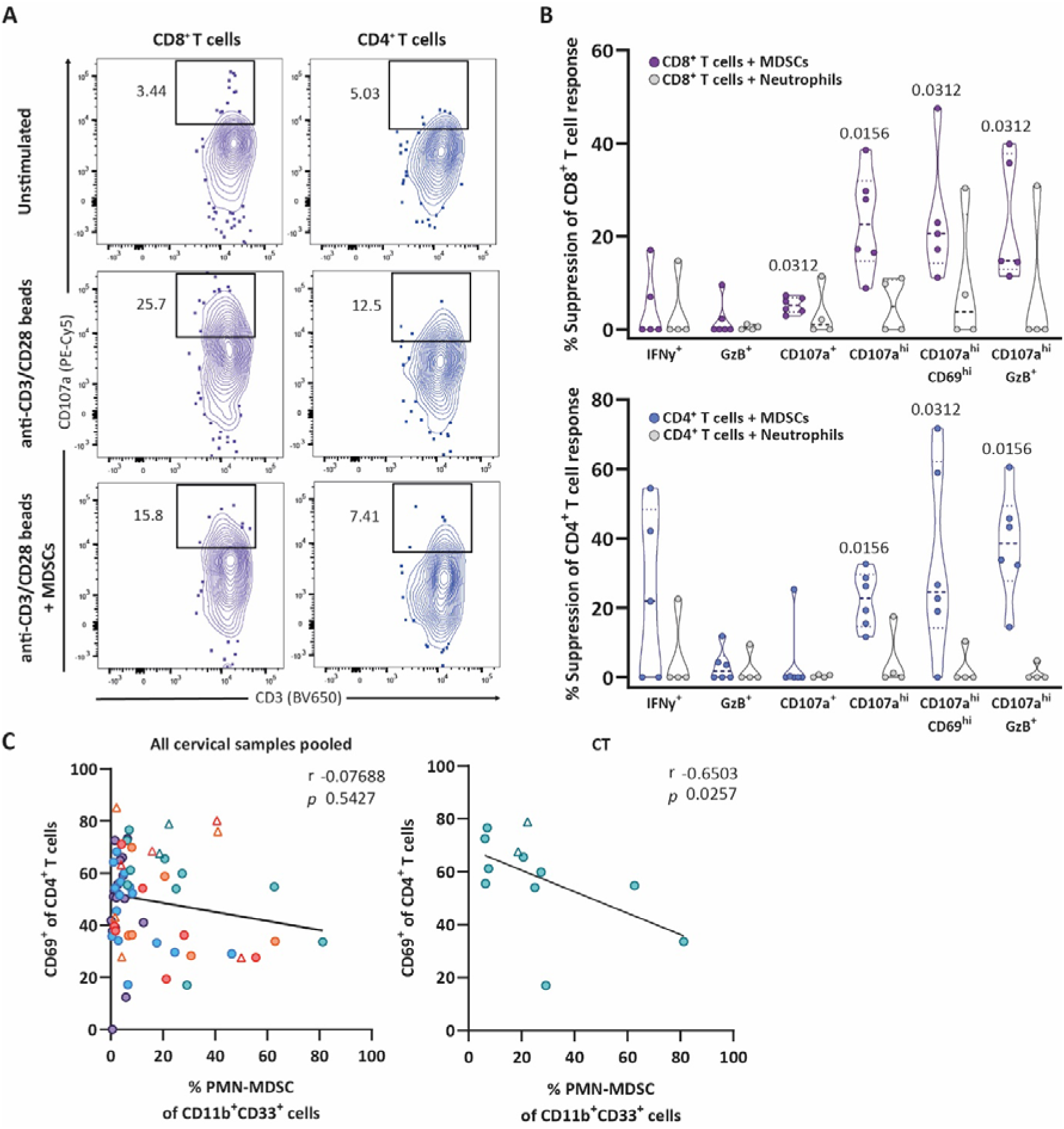
Cervical MDSCs suppress T-cell activation and cytotoxic potential. (A) Representative flow-cytometry plots showing CD8^+^ and CD4^+^ T cells and their expression of degranulation marker CD107a after exposure to several conditions for 24 hours: unstimulated, anti- CD3/anti-CD28 beads stimulated, and anti-CD3/anti-CD28 beads stimulated in the presence of MDSC cells (1:5 MDSC to lymphocyte ratio) or control neutrophils (1:5 neutrophil to lymphocyte ratio). (B) Calculated percentage of suppression mediated by MDSCs (n=6) or neutrophils (n=4) of the indicated markers expressed by CD8^+^ (purple) and CD4^+^ (blue) T cells. (C) Graphs show the relationship between the frequency of CD69^+^ CD4^+^ T cells and the frequency of PMN-MDSCs found in the cervix of all included women (left) and of CT patients only (right). Data are shown as violin plots with median and quartiles (B). Patients with leukocyte/HPF counts of >10 are indicated with triangles. Statistical significance was determined by Wilcoxon test (one-sided) (B). Correlations (*r* and *P* values) were assessed by Spearman test (two-sided) (C).

Last, to translate these findings to the patients included in our study, we assessed associations between the presence of PMN- and M-MDSCs in the cervical mucosa with the frequency of CD69^+^ T cells, indicative of T cell activation and/or tissue residence (35, 36). Amongst all women included in our study we did not find an association between the frequency of cervical mucosa PMN-MDSCs and CD69^+^ CD4^+^ T cells (**Figure 6C**). However, in CT patients, we observed a negative correlation between the frequency of cervical mucosa PMN-MDSCs and CD69^+^ CD4^+^ T cells (**Figure 6C**). Together, these data confirm that MDSCs residing in cervical mucosa are capable of suppressing antimicrobial effector functions of cervical CD4^+^ and CD8^+^ T cells.

## Discussion

In this study we identified an expansion of PMN-MDSCs in the genital tract of women with CT infection, BV, and coinfections, which is mediated by signals in the mucosal milieu, including the inflammatory cytokine IL-1β. Furthermore, these local signals induced MDSC subsets to express the immune suppressive molecules PD-L1 and IDO (37, 38). Using a cervical explant model, we confirm the link between CT exposure, IL-1β secretion and potentially suppressive PMN-MDSCs expansion. Last, we demonstrate that cervical MDSC limit activation and function of cervical T cells, all together likely reducing time to pathogen clearance while potentially enhancing vulnerability to secondary infections at this site and compromising overall women’s health.

Increased levels of PMN-MDSCs have previously been described in cancer, autoimmune disease (15, 16), COVID-19 (34, 39), and chronic viral infections such as HIV (24, 40). However, to date, the role of MDSCs in the context of localized genital tract conditions in women remained unknown. In mice, a recent report showed that *in vivo* infection with CT increased levels of MDSCs in the cervix, which could be reversed by inhibition of the yes-associated protein 1 (41). However, due to a limited set of phenotypical markers used, subsets of MDSCs were not identified (41). We now show elevated PMN-MDSC levels in the cervical mucosa of women with CT, as well as with BV or a coinfection, but not in women with HPV. Indeed, MDSCs have been highlighted as a mechanism limiting cervical tumor clearance (42) and HPV patients have been shown to accumulate MDSCs in the tumor (43). Yet, women included in the HPV group of our study were diagnosed with low- or high-grade squamous intraepithelial lesions, but not with cervical cancer, potentially explaining unaltered levels of MDSCs compared to the control group.

The levels of PMN-MDSCs remained similar in blood across all patient groups, which reinforces the importance of assessing the local immune environment in compartmentalized mucosal- and tissue-specific conditions. Indeed, not all infections elicit similar mucosal versus systemic MDSCs alterations, as M-MDSCs and PMN-MDSCs have previously been shown to increase in blood of COVID-19 patients but not in the airway mucosa, while for influenza-infected patients mucosal levels where higher compared to blood (34). In this sense, the level of localized versus systemic inflammation and sample timing may have an impact on these findings.

CT infection often presents as a silent, asymptomatic infection in women (44), which is associated with CT serovars D and I (45). This may explain why CVL fluid cytokine levels from asymptomatic CT patients included in our study showed minimal differences from those in HD, unlike the CVL fluid profiles observed in symptomatic BV and coinfected patients. Nonetheless, we detected a positive correlation between cervical mucosal IL-1β levels and cervical PMN-MDSC frequencies in these CT patients, indicating a potential role for IL-1β in PMN-MDSC expansion. Indeed, IL-1β has been reported as a driver of suppressive PMN-MDSC expansion in tumor-bearing mice (46, 47), through activation of extracellular signal-regulated kinases 1 and 2 (48). Moreover, levels of IL-1β positively correlated with PMN-MDSC frequencies in the blood of COVID-19 patients (49) and IL-1β has been described as a potential inducer of PD-L1 and IDO expression in myeloid cells (50, 51), which we indeed partially observed in both cervical PMN- and M-MDSCs. These findings warrant further exploration of blocking IL-1β and/or signaling pathways to reduce PMN-MDSC levels and activity.

Patients with BV and Coinf showed elevated levels of several cytokines in the CVL compared to HD. Levels of VEGF-A were higher in BV CVL and correlated with increased levels of mucosal PMN-MDSCs. VEGF may function as a chemoattractant for MDSCs (52), which is further propagated by MDSC production of matrix metalloproteinase 9 and may allow the release of additional VEGF into the micro-environment (53). Moreover, we found that exposure of cervical tissue to CVL from BV patients elevated PMN-MDSC frequencies and IDO expression, suggesting a role for the milieu signature associated to these samples in PMN-MDSC expansion and activity. Overall, other factors present in the cervical mucosal micro-environment may contribute to expand PMN-MDSCs and/or modify their function (15). In this sense, the local cytokine micro-environment and thus the level of PMN-MDSCs in the cervical mucosa may be determined by the composition of genital bacterial communities (5), of which the complexity is specifically increased due to an overgrowth of anaerobic bacteria during BV, such as *Gardnerella vaginalis* (54).

By isolating cervical tissue MDSCs we were able to demonstrate the suppressive capacity of these tissue-resident MDSCs. We showed that MDSCs suppressed degranulation and cytotoxic factors of CD4^+^ and CD8^+^ T cells are required for optimal pathogen clearance (27, 28), suggesting that MDSCs found in patients associated to disturbances of the FGT microenvironment are likely suppressive. As induction of IDO and PD-L1 enhances the immune suppressive capacity of MDSCs (37, 38), our results indicate that, while only PMN-MDSCs are expanded after EB or CT CVL stimulation, both PMN-and M-MDSCs may increase their suppressive capacity as a consequence of exposure to CT. Future research should separately assess suppression by PMN-MDSC and M-MDSCs, which was limited in the current study due to low sorted cell numbers acquired from cervical samples. Single cell analyses could offer insight in the functional diversity of these cells yet variability between patients may complicate conclusions. Nonetheless, our results indicate an effect of FGT conditions on the suppressive capacity of both PMN- and M-MDSC subsets and warrant further exploration of reducing cervical MDSC levels and blocking suppressive mechanisms, such as PD-L1.

Whether age has an impact on PMN-MDSC frequencies in the cervical mucosa remains to be elucidated. Expansion of MDSCs has been reported to occur during aging in the blood and secondary lymphoid organs of mice (55), which is mainly explained by a chronic low-grade increase in levels of inflammatory cytokines during aging, including cytokines reported to promote MDSC expansion (56). Moreover, higher MDSC levels in the blood of COVID-19 patients have recently been shown to be associated with age (34). As age differed between our patient groups, with younger ages present in the CT and Coinf groups, our findings indicate that higher levels of PMN-MDSCs in the groups with younger age were most likely caused by these conditions of the cervical mucosa rather than an age-related increase of PMN-MDSCs. Furthermore, cervical MDSCs may expand due to cyclic changes caused by estrogens (57) and pregnancy (18). Whereas the menstrual cycle stage differed between groups, we found that this difference did not appear to significantly change mucosal MDSC frequencies nor CVL fluid cytokine levels. Differences in PMN-MDSC levels were maintained when the two dominant phases of the menstrual cycle, follicular and luteal, were separated (58).We did not observe major changes to MDSC levels and CLV fluid cytokine levels as a consequence of hormonal contraceptives. However, as hormonal contraceptives may influence MDSC levels (59), we were not able to definitively exclude an effect of hormone-induced alterations to MDSC levels in our study cohort.

Differentiation and recruitment of MDSCs during conditions of the FGT has likely evolved to prevent over-activation of effector cells and thereby tissue pathology. However, enhanced immune suppression may also negatively impact effector responses to pathogens, resulting in less efficient immune clearance while likely increase vulnerability to secondary STIs such as HIV. Our findings provide a starting point for designing preventive and therapeutic strategies limiting MDSC expansion induced by inflammatory mediators, which could improve natural pathogen clearance, potentially lowering antibiotic usage and resistance, and limiting vulnerability to secondary STIs. Altogether, we identified unique features of resident immune cells influenced by the local mucosal microenvironment crucial for enhancing mucosal immunity and ensuring women’s sexual and reproductive health.

## Methods

### Ethics statement

Written informed consent was obtained from all participants prior to inclusion into this study. This study was performed in accordance with the Declaration of Helsinki and approved by the corresponding Institutional Review Board (PR(AG)117/2018) of the Vall d’Hebron University Hospital (HUVH).

### Study Design

Women visiting the Drassanes Vall d’Hebron Centre for International Health and Infectious Diseases in Barcelona, Spain due to a self-suspected FGT condition were included in this study and allocated to their study group after laboratory confirmed diagnosis. All participants assigned to the CT, BV and coinfection groups were tested for presence of several pathogens. Presence of *C. trachomatis*, *Mycoplasma genitalium*, *Mycoplasma hominis*, *Neisseria gonorrhoeae*, *Trichomonas vaginalis*, *Ureaplasma parvum*, and *Ureaplama urealyticum* was determined by multiplex nucleic acid amplification test (NAAT) (Allplex^TM^ STI Essential Assay, Seegene Inc.) according to the manufacturer’s recommendations. For the diagnosis of bacterial vaginosis, presence of grade 3 of the Ison-Hay criteria of the corresponding vaginal Gram stain was used (60). Additionally, in all samples, presence of *Candida spp*. was determined using Candida BBL chromogenic plates (BD), which were incubated at 37°C for a minimum of 48 hours (61). Participants who were solely positive for *C. trachomatis* and presented with an asymptomatic infection were included in the CT group, while participants with a grade 3 ISON-Hay score and negative for other pathogens were assigned to the BV group. The coinfection group mainly consisted of participants with CT and BV conditions (**Supplemental Table 2** for specifics). All participants were tested negative for *T. vaginalis*. Additionally, to estimate aerobic vaginitis (AV), also known as desquamative inflammatory vaginitis, and considering that fresh examination using phase-contrast microscopy was not available in our laboratory, suspected cases would be based on grade 4 according to the Ison-Hay criteria together with >10 leukocytes per field in Gram staining, according to current protocols in our laboratory. While these has obvious limitations, since our protocol does not include the evaluation of parabasal cells, which limits diagnostic accuracy (62), no sample included in this study scored grade 4 based on Ison- Hay criteria.

In addition, women allocated to the HPV group were diagnosed with HPV infection using the Hybrid Capture 2 system (Qiagen), followed by the CLART-HPV2 test (Genomica, Madrid, Spain) (63), which is a polymerase chain reaction technique that allows for the detection of 35 HPV genotypes, or using the Cobas HPV Test (Roche Molecular System). Women without STI or conditions of the FGT were recruited at the Gynecology and Obstetrics department of the HUVH, Barcelona, Spain. Exclusion criteria were: HIV or hepatitis, immune-related illnesses, systemic illnesses, pregnancy, and menstruation at the time of visit. All women were asked to fill in a questionnaire, registering prior altered FGT conditions and other immunological or allergic conditions, medication, menstrual cycle stage at time of the study and use of hormonal contraceptives. Cervical tissue used in this study was obtained from patients, aged 41-82 years (median age 60 years), who were recruited at the Gynecology Unit of the HUVH and were undergoing a partial or radical hysterectomy for non-tumoral and non-inflammatory indications. Low numbers of cellular subsets (<100 cells) in clinical samples detected by flow cytometry were excluded. For cytokine analyses, extreme outliers were excluded based on the ROUT method (Q=0.1% to remove definitive outliers) in Prism 8.3.0 (GraphPad Software, La Jolla, CA, USA).

### Sample collection

Whole blood from women with and without FGT conditions was collected in EDTA-containing tubes. Plasma was isolated from these samples and stored at −80°C. Subsequently, PBMCs were isolated via Ficoll–Paque separation. Part of these PBMCs were immediately used for flow cytometry analyses, while the remainder was stored in liquid nitrogen. From the same women, an endocervical sample was collected with a cytobrush and cervicovaginal lavage (CVL) fluid was obtained by introducing 5mL of physiological serum into the cervicovaginal canal and the posterior fornix, after which the serum was retrieved again. Cervical samples were kept at 4°C and were sent to the laboratory for immediate processing (<4 hours after collection). Cervical cytobrush samples were vortexed, washed with PBS, filtered through a 100µm pore-size cell strainer (Labclinics), and centrifuged, before immediate labelling for flow cytometry. Of note, we did not perform cell separation methods of cytobrush samples due to low cell numbers. CVL fluid samples were centrifuged to remove mucus and debris. Subsequently, the supernatant was stored at −80°C, while remaining cells in the cell pellet were added to the cervical cytobrush samples.

Endo- and ectocervical tissue was collected from patients at the HUVH undergoing partial or radical hysterectomy and stored in antibiotic-containing RPMI 1640 medium, after pathological inspection. Endo- and ectocervical tissue was immediately dissected into 8-mm^3^ tissue blocks as reported previously (36, 64). Depending on the assay, tissue blocks were either immediately enzymatically digested or first cultured in several conditions described below before tissue digestion. Enzymatic digestion was initiated by adding 10 mg/mL collagenase IV (Gibco) in RPMI +5% FBS to the tissue blocks, incubated for 30 mins at 37°C and 400 rpm before mechanically digestion with a pestle (36, 64). Cell suspensions were filtered through a 70µm pore-size cell strainer (Labclinics) and prepared for flow cytometry as described below.

### Flow cytometry of patient PBMC and Cytobrush

Single cell suspensions of PBMC and cytobrush samples were stained with Live/Dead Aqua (Invitrogen) before extracellular labeling with antibodies targeting the following proteins: CD33 (PerCP-Cy5.5, Clone WM53, Biolegend, #303414), CD11b (FITC, Clone M1/70, Biolegend, #101205), CD3 (PE-Cy7, Clone SK7, BD Biosciences, #557851), CD69 (PE-CF594, Clone FN50, BD Biosciences, #562617), CD56 (PE, Clone B159, BD Biosciences, #555516), CD14 (APC-H7, Clone MφP9, BD Biosciences, #560180), CD45 (AF700, Clone HI30, Biolegend, #304024), CD8 (APC, Clone RPA-T8, BD Biosciences, #561952), CD16 (BV786, Clone 3G8, BD Biosciences, #563690), CD103 (BV650, Clone Ber-ACT8, BD Biosciences, #743653), CD15 (BV605, Clone W6D3, BD Biosciences, #562980), and HLA-DR (BV421, Clone G46-6, BD Biosciences, #562804). Labeling was performed at room temperature for 20 mins, washed with PBS +3% FBS (PBMCs) or washed with PBS +3% normal mouse serum (NMS) and normal goat serum (NGS), and fixed with PBS+1% paraformaldehyde before acquisition on a BD LSR Fortessa flow cytometer (Cytomics Platform, High Technology Unit (UAT), Vall d’Hebron Institut de Recerca (VHIR)).

### Measurement of cytokines

Cytokines present in plasma and CVL fluid of FGT inflammatory patients as well as in *ex vivo* cervical tissue supernatants were detected using an automated multiplex ELISA system (Ella Simple Plex™, Bio-Techne R&D Systems). Levels of the following eight analytes were detected in these samples according to the manufacturer’s instructions: CSF1, G-CSF, GM-CSF, VEGF-A, IL-1β, IL-6, TNF-α, TGF-β1. Levels of analytes found in CVL fluid samples were normalized to the total amount of protein found in CVL fluid as we did not obtain equal volumes of CVL fluid across all patients. Total protein content was measured using the Atellica CH automated equipment (Siemens Healthineers®) with a colorimetric methodology based on the biuret test.

### Flow cytometry of cervical tissue

After digestion of cervical tissue blocks and depending on the assay, single cell suspensions were stained with Live/Dead Aqua (Invitrogen) before extracellular labeling with antibodies targeting the following proteins: HLA-DR (PerCP-Cy5.5, Clone G46-6, BD Biosciences, #560652), CD11b (FITC, Clone M1/70, Biolegend, #101205), PD-L1 (PE-Cy7, Clone 29E.2A3, Biolegend, #329718), CD14 (APC-H7, Clone MφP9, BD Biosciences, #560180), CD45 (AF700, Clone HI30, Biolegend, #304024), CD15 (BV605, Clone W6D3, BD Biosciences, #562980), and CD33 (BV421, Clone P67.6, Biolegend, #366622). Extracellular labeling was performed at room temperature for 20 minutes in PBS+ 3% NMS/NGS and washed with PBS +3% NMS/NGS. For intracellular antibody labeling, cells were fixed for 20 mins at RT (Medium A, Invitrogen) and labeled with antibodies targeting IDO (PE,

Clone eyedio, Invitrogen, #12-9477-42) in permeabilizing medium (Medium B, Invitrogen) for 30 minutes at RT. Cells were then washed with PBS +3% NMS/NGS, and fixed with PBS+1% paraformaldehyde before acquisition on a BD LSR Fortessa flow cytometer (Cytomics Platform, UAT, VHIR).

### Purification of Chlamydia trachomatis elementary bodies and infection of cervical tissue

HeLa-229 cells (ATCC CCL-2.1) were infected with *C. trachomatis* Serovar D (ATCC VR-885) as previously described (65), with minor modifications. Briefly, HeLa 229 cells were maintained in DMEM (+10% FBS + penicillin/streptomycin) at 37°C before chlamydia infection. To infect HeLa cells, confluent cell monolayers grown in 25cm^2^ flasks (6x10^6^ cells/well) and exposed to thawed *C. trachomatis* elementary bodies (EBs) in DMEM (+10% FBS +10µg/mL gentamycin +1µg/mL cycloheximide). Flasks were centrifuged (30 mins at 754 x *g*) and placed at 37°C. After 48h of incubation and/or observation of the characteristic cytopathic effect (cytoplasmic inclusion bodies) in 90-100% of the infected cells, cells were collected into tubes by scraping and centrifuged at 400 x *g* for 7 mins to remove the supernatant. The cell pellet was resuspended in ice cold PBS, transferred to a bijou tube with 5-7 glass beads (Thermo Fisher, #11369123) and briefly vortexed. After centrifugation, the supernatant was mixed 1:1 with sucrose-phosphate-glutamate buffer (SPG) and stored at -80°C until further use. To titrate the infective stock, 1:10 serial dilutions were made in HeLa229 cells plated in a 96-well plate. After 48-72 h of incubation at 37°C and 5% CO2, infected cell monolayers were washed twice with PBS and fixed in 96% ice cold methanol for 10 min at -20°C. After washing and blocking unspecific unions with 3% BSA-PBS at RT for 60min, chlamydial inclusions were stained with FITC-conjugated anti-Chlamydia antibody (Chemicon #AB1120F; 1/200 dilution t RT for) at RT for 1h. Inclusions were counted by observation with a fluorescence microscope and results were expressed in inclusion forming units (IFU/ml).

For tissue infection, fresh ecto- and endocervical tissue blocks (8 mm^3^) were placed in a 24 wells plate with the mucosa facing upwards and exposed to RPMI 1640 (+20% FBS, without penicillin) alone, or RPMI 1640 (+20% FBS) with *C. trachomatis* EB (1*10^5^ IFU/mL) or GM-CSF (10 ng/mL) and IL-6 (10 ng/mL) directly placed into the mucosal interface and incubated for 48 hours at 37°C After incubation, tissue blocks were digested and prepared for flow cytometry to assess MDSC frequencies and phenotype as described above.

### Exposure of cervical tissue to CVL fluid

Cervical tissue blocks (8 mm^3^) were placed in a 24-well plate in the presence of equal amounts of RPMI 1640 + 20% FBS and CVL fluid of CT, BV, or HD patients. Before use, CVL fluid samples were thawed and centrifuged to get rid of potential debris. Additionally, cervical tissue was exposed to IL-1β (100 ng/mL). Cervical tissue blocks were then incubated for 24 hours at 37°C. After incubation, tissue blocks were digested and prepared for flow cytometry to assess MDSC frequencies and phenotype, including intracellular markers, as described above.

### MDSC suppression assay

Single cell suspensions of freshly obtained cervical tissue were stained with Live/Dead Aqua (Invitrogen) and extracellularly labeled with antibodies targeting the following proteins: HLA-DR (PerCP-Cy5.5, Clone G46-6, BD Biosciences, #560652), CD11b (FITC, Clone M1/70, Biolegend, #101205), CD14 (APC-H7, Clone MφP9, BD Biosciences, #560180), CD45 (BV605, clone HI30, BD Biosciences, #564047), CD20 (V500, Clone, BD Biosciences, #647463), and CD33 (BV421, Clone P67.6, Biolegend, #366622). Cell suspensions were then resuspended in PBS+3% FBS and sorted into lymphocyte (based on FSC and SSC size) and MDSC (CD14^+^ and CD14^-^ HLA-DR^low^ cells of CD33^+^CD11b^+^ myeloid cells) fractions using the Cytek Aurora Cell Sorter (Cytek Biosciences). Suppression assays were performed by culturing duplicates or triplicates of sorted lymphocytes with and without MDSCs in a ratio of 1:5 (MDSC:lymphocyte) in U-bottom 96-wells plates in the presence or absence of anti-CD3/anti-CD28 coupled beads (1:2 bead-to-lymphocyte ratio) (Dynabeads™, Invitrogen), CD107a (PE-Cy5, clone H4A3, BD Biosciences, #555802), brefeldin A (0.55_μL/mL), monensin (0.385_μL/mL) for 18 hours at 37°C. After culturing, duplicates or triplicates were pooled, washed with PBS, and fixed and permeabilized for intracellular labeling (Foxp3/Transcription Factor Staining Buffer, eBioscience) with the following antibodies: CD69 (PE-CF594, clone FN50, BD Biosciences, #562617), Granzyme B (PE, Clone GB11, Thermo Fisher, #GRB04), IFN-γ (AF700, Clone B27, Invitrogen, #MHCIFG29), CD8 (APC, Clone RPA-T8, BD Biosciences, #561952), and CD3 (BV650, Clone UCHT1, BD Biosciences, #563851). Cells were acquired on a BD LSR Fortessa flow cytometer (Cytomics Platform, UAT, VHIR).

### Statistics

Data shown in bar graphs were expressed as median and interquartile range. Two-sided Wilcoxon matched-pairs signed rank test was applied for paired comparisons, while a one-sided test was applied to fold change data. Kruskal–Wallis rank–sum test with Dunn’s post hoc test was used for multiple comparisons. Correlation analyses were performed using non-parametric Spearman rank correlation. Linear regression analysis was performed to generate lines of best fit. Suppression in suppression assays were calculated as follows: ((% marker T cell only – % marker T cell with MDSC suppressor cells) / (% marker T cell only)) × 100%. Statistical significance was then determined by one-sided Wilcoxon matched-pairs singed rank test by comparing to the control. A *P* value < 0.05 was considered statistically significant. No statistical method was used to predetermine sample size, as this was dependent on patient consent and eligibility to the study groups. Flow-cytometry data were analyzed using FlowJo v10.7.1 software (TreeStar). Data and statistical analyses were performed using Prism 8.3.0 (GraphPad Software, La Jolla, CA, USA).

## Data availability

All data are available in the main text or the supplemental materials. Source data are provided with this paper.

## Acknowledgements

We would like to thank all the patients who participated in the study and Joan Puñet Ortiz and Sara Monreal Peinado for assistance with cell sorting at the Cytomics Platform, UAT, VHIR. Figure 1A was created with BioRender.com. This work was supported by grants from Fundació La Marató TV3 (FMTV3) 201814-10 (MG) and 202112-30 (MG), the Spanish Health Institute Carlos III (ISCIII) co- funded by ERDF/ESF, “A way to make Europe”/“Investing in your future” PI20/00160 (MG), Gilead fellowship GLD18/00008 (MG), the Spanish Health Institute Carlos III, Miguel Servet program CP17/00179 (MJB), and Vall d’Hebron Institut de Recerca (VHIR) PhD Fellowship (NM). The funders had no role in study design, data collection and analysis, the decision to publish, or the preparation of the manuscript.

## Author contributions

Conceptualization, MG; Methodology, DKJP, ABM, NM, JC, YHM, DAS, CR; Patient recruitment and sample collection, VD, MA, PA, JNG, LMB, CCM, NLR, JC, VF; Investigation, DKJP, ABM, NM, JC, MJB; Formal analysis, DKJP, NM, JC, MJB, MG; Supervision, MG; Writing original draft, DKJP, MG; Writing review & editing, all authors.

## References

1. (WHO) WHO. Sexually transmitted infections (STIs) (Fact sheet). https://www.who.int/news-room/fact-sheets/detail/sexually-transmitted-infections-(stis). Accessed 08-03-2024, 2024.

2. (WHO) WHO. Bacterial vaginosis (Fact sheet). https://www.who.int/news-room/fact-sheets/detail/bacterial-vaginosis. Accessed 8-03-2024, 2024.

3. Chan D, Bennett PR, Lee YS, Kundu S, Teoh TG, Adan M, et al. Microbial-driven preterm labour involves crosstalk between the innate and adaptive immune response. Nat Commun. 2022;13(1):975.

4. Atashili J, Poole C, Ndumbe PM, Adimora AA, and Smith JS. Bacterial vaginosis and HIV acquisition: a meta-analysis of published studies. AIDS. 2008;22(12):1493–501.

5. Gosmann C, Anahtar MN, Handley SA, Farcasanu M, Abu-Ali G, Bowman BA, et al. Lactobacillus-Deficient Cervicovaginal Bacterial Communities Are Associated with Increased HIV Acquisition in Young South African Women. Immunity. 2017;46(1):29–37.

6. van Teijlingen NH, Helgers LC, Sarrami-Forooshani R, Zijlstra-Willems EM, van Hamme JL, Segui-Perez C, et al. Vaginal bacterium Prevotella timonensis turns protective Langerhans cells into HIV-1 reservoirs for virus dissemination. EMBO J. 2022;41(19):e110629.

7. Buckner LR, Amedee AM, Albritton HL, Kozlowski PA, Lacour N, McGowin CL, et al. Chlamydia trachomatis Infection of Endocervical Epithelial Cells Enhances Early HIV Transmission Events. PLoS One. 2016;11(1):e0146663.

8. Liebenberg LJP, McKinnon LR, Yende-Zuma N, Garrett N, Baxter C, Kharsany ABM, et al. HPV infection and the genital cytokine milieu in women at high risk of HIV acquisition. Nat Commun. 2019;10(1):5227.

9. Houlihan CF, Larke NL, Watson-Jones D, Smith-McCune KK, Shiboski S, Gravitt PE, et al. Human papillomavirus infection and increased risk of HIV acquisition. A systematic review and meta-analysis. AIDS. 2012;26(17):2211–22.

10. Mlisana K, Naicker N, Werner L, Roberts L, van Loggerenberg F, Baxter C, et al. Symptomatic vaginal discharge is a poor predictor of sexually transmitted infections and genital tract inflammation in high-risk women in South Africa. J Infect Dis. 2012;206(1):6–14.

11. Haaland RE, Hawkins PA, Salazar-Gonzalez J, Johnson A, Tichacek A, Karita E, et al. Inflammatory genital infections mitigate a severe genetic bottleneck in heterosexual transmission of subtype A and C HIV-1. PLoS Pathog. 2009;5(1):e1000274.

12. Ghys PD, Fransen K, Diallo MO, Ettiegne-Traore V, Coulibaly IM, Yeboue KM, et al. The associations between cervicovaginal HIV shedding, sexually transmitted diseases and immunosuppression in female sex workers in Abidjan, Cote d’Ivoire. AIDS. 1997;11(12):F85–93.

13. Laga M, Manoka A, Kivuvu M, Malele B, Tuliza M, Nzila N, et al. Non-ulcerative sexually transmitted diseases as risk factors for HIV-1 transmission in women: results from a cohort study. AIDS. 1993;7(1):95–102.

14. Johnson LF, and Lewis DA. The effect of genital tract infections on HIV-1 shedding in the genital tract: a systematic review and meta-analysis. Sex Transm Dis. 2008;35(11):946–59.

15. Veglia F, Sanseviero E, and Gabrilovich DI. Myeloid-derived suppressor cells in the era of increasing myeloid cell diversity. Nat Rev Immunol. 2021;21(8):485–98.

16. Park SJ, Nam DE, Seong HC, and Hahn YS. New Discovery of Myeloid-Derived Suppressor Cell’s Tale on Viral Infection and COVID-19. Front Immunol. 2022;13:842535.

17. Medina E, and Hartl D. Myeloid-Derived Suppressor Cells in Infection: A General Overview. J Innate Immun. 2018;10(5-6):407–13.

18. Kostlin N, Kugel H, Spring B, Leiber A, Marme A, Henes M, et al. Granulocytic myeloid derived suppressor cells expand in human pregnancy and modulate T-cell responses. Eur J Immunol. 2014;44(9):2582–91.

19. Pan T, Liu Y, Zhong LM, Shi MH, Duan XB, Wu K, et al. Myeloid-derived suppressor cells are essential for maintaining feto-maternal immunotolerance via STAT3 signaling in mice. J Leukoc Biol. 2016;100(3):499–511.

20. Liebenberg LJ, Masson L, Arnold KB, McKinnon LR, Werner L, Proctor E, et al. Genital- Systemic Chemokine Gradients and the Risk of HIV Acquisition in Women. J Acquir Immune Defic Syndr. 2017;74(3):318–25.

21. Masson L, Arnold KB, Little F, Mlisana K, Lewis DA, Mkhize N, et al. Inflammatory cytokine biomarkers to identify women with asymptomatic sexually transmitted infections and bacterial vaginosis who are at high risk of HIV infection. Sex Transm Infect. 2016;92(3):186–93.

22. Masson L, Passmore JA, Liebenberg LJ, Werner L, Baxter C, Arnold KB, et al. Genital inflammation and the risk of HIV acquisition in women. Clin Infect Dis. 2015;61(2):260–9.

23. Bronte V, Brandau S, Chen SH, Colombo MP, Frey AB, Greten TF, et al. Recommendations for myeloid-derived suppressor cell nomenclature and characterization standards. Nat Commun. 2016;7:12150.

24. Wang L, Zhao J, Ren JP, Wu XY, Morrison ZD, Elgazzar MA, et al. Expansion of myeloid-derived suppressor cells promotes differentiation of regulatory T cells in HIV-1+ individuals. AIDS. 2016;30(10):1521–31.

25. Sacchi A, Tumino N, Sabatini A, Cimini E, Casetti R, Bordoni V, et al. Myeloid-Derived Suppressor Cells Specifically Suppress IFN-gamma Production and Antitumor Cytotoxic Activity of Vdelta2 T Cells. Front Immunol. 2018;9:1271.

26. Tumino N, Besi F, Martini S, Di Pace AL, Munari E, Quatrini L, et al. Polymorphonuclear Myeloid-Derived Suppressor Cells Are Abundant in Peripheral Blood of Cancer Patients and Suppress Natural Killer Cell Anti-Tumor Activity. Front Immunol. 2021;12:803014.

27. Lampe MF, Wilson CB, Bevan MJ, and Starnbach MN. Gamma interferon production by cytotoxic T lymphocytes is required for resolution of Chlamydia trachomatis infection. Infect Immun. 1998;66(11):5457–61.

28. Fankhauser SC, and Starnbach MN. PD-L1 limits the mucosal CD8+ T cell response to Chlamydia trachomatis. J Immunol. 2014;192(3):1079–90.

29. Russell AN, Zheng X, O’Connell CM, Wiesenfeld HC, Hillier SL, Taylor BD, et al. Identification of Chlamydia trachomatis Antigens Recognized by T Cells From Highly Exposed Women Who Limit or Resist Genital Tract Infection. J Infect Dis. 2016;214(12):1884–92.

30. Gondek DC, Roan NR, and Starnbach MN. T cell responses in the absence of IFN-gamma exacerbate uterine infection with Chlamydia trachomatis. J Immunol. 2009;183(2):1313–9.

31. Stary G, Olive A, Radovic-Moreno AF, Gondek D, Alvarez D, Basto PA, et al. VACCINES. A mucosal vaccine against Chlamydia trachomatis generates two waves of protective memory T cells. Science. 2015;348(6241):aaa8205.

32. Schust DJ, Ibana JA, Buckner LR, Ficarra M, Sugimoto J, Amedee AM, et al. Potential mechanisms for increased HIV-1 transmission across the endocervical epithelium during C. trachomatis infection. Curr HIV Res. 2012;10(3):218–27.

33. Negorev D, Beier UH, Zhang T, Quatromoni JG, Bhojnagarwala P, Albelda SM, et al. Human neutrophils can mimic myeloid-derived suppressor cells (PMN-MDSC) and suppress microbead or lectin-induced T cell proliferation through artefactual mechanisms. Sci Rep. 2018;8(1):3135.

34. Falck-Jones S, Vangeti S, Yu M, Falck-Jones R, Cagigi A, Badolati I, et al. Functional monocytic myeloid-derived suppressor cells increase in blood but not airways and predict COVID-19 severity. J Clin Invest. 2021;131(6).

35. Kumar BV, Ma W, Miron M, Granot T, Guyer RS, Carpenter DJ, et al. Human Tissue-Resident Memory T Cells Are Defined by Core Transcriptional and Functional Signatures in Lymphoid and Mucosal Sites. Cell Rep. 2017;20(12):2921–34.

36. Cantero-Perez J, Grau-Exposito J, Serra-Peinado C, Rosero DA, Luque-Ballesteros L, Astorga-Gamaza A, et al. Resident memory T cells are a cellular reservoir for HIV in the cervical mucosa. Nat Commun. 2019;10(1):4739.

37. Noman MZ, Desantis G, Janji B, Hasmim M, Karray S, Dessen P, et al. PD-L1 is a novel direct target of HIF-1alpha, and its blockade under hypoxia enhanced MDSC-mediated T cell activation. J Exp Med. 2014;211(5):781–90.

38. Yu J, Du W, Yan F, Wang Y, Li H, Cao S, et al. Myeloid-derived suppressor cells suppress antitumor immune responses through IDO expression and correlate with lymph node metastasis in patients with breast cancer. J Immunol. 2013;190(7):3783–97.

39. Agrati C, Sacchi A, Bordoni V, Cimini E, Notari S, Grassi G, et al. Expansion of myeloid-derived suppressor cells in patients with severe coronavirus disease (COVID-19). Cell Death Differ. 2020;27(11):3196–207.

40. Vollbrecht T, Stirner R, Tufman A, Roider J, Huber RM, Bogner JR, et al. Chronic progressive HIV-1 infection is associated with elevated levels of myeloid-derived suppressor cells. AIDS. 2012;26(12):F31–7.

41. Lu X, Wang Y, Ma Y, Huang D, Lu Y, Liu X, et al. YAP1 induces marrow derived suppressor cell recruitment in Chlamydia trachomatis infection. Immunol Lett. 2022;242:8–16.

42. Barros MR, Jr., de Melo CML, Barros M, de Cassia Pereira de Lima R, de Freitas AC, and Venuti A. Activities of stromal and immune cells in HPV-related cancers. J Exp Clin Cancer Res. 2018;37(1):137.

43. Liang Y, Wang W, Zhu X, Yu M, and Zhou C. Inhibition of myeloid-derived suppressive cell function with all-trans retinoic acid enhanced anti-PD-L1 efficacy in cervical cancer. Sci Rep. 2022;12(1):9619.

44. Witkin SS, Minis E, Athanasiou A, Leizer J, and Linhares IM. Chlamydia trachomatis: the Persistent Pathogen. Clin Vaccine Immunol. 2017;24(10).

45. Lan J, Melgers I, Meijer CJ, Walboomers JM, Roosendaal R, Burger C, et al. Prevalence and serovar distribution of asymptomatic cervical Chlamydia trachomatis infections as determined by highly sensitive PCR. J Clin Microbiol. 1995;33(12):3194–7.

46. Elkabets M, Ribeiro VS, Dinarello CA, Ostrand-Rosenberg S, Di Santo JP, Apte RN, et al. IL- 1beta regulates a novel myeloid-derived suppressor cell subset that impairs NK cell development and function. Eur J Immunol. 2010;40(12):3347–57.

47. Fridlender ZG, Sun J, Mishalian I, Singhal S, Cheng G, Kapoor V, et al. Transcriptomic analysis comparing tumor-associated neutrophils with granulocytic myeloid-derived suppressor cells and normal neutrophils. PLoS One. 2012;7(2):e31524.

48. Shi H, Qin Y, Tian Y, Wang J, Wang Y, Wang Z, et al. Interleukin-1beta triggers the expansion of circulating granulocytic myeloid-derived suppressor cell subset dependent on Erk1/2 activation. Immunobiology. 2022;227(1):152165.

49. Sacchi A, Grassi G, Bordoni V, Lorenzini P, Cimini E, Casetti R, et al. Early expansion of myeloid-derived suppressor cells inhibits SARS-CoV-2 specific T-cell response and may predict fatal COVID-19 outcome. Cell Death Dis. 2020;11(10):921.

50. Karakhanova S, Meisel S, Ring S, Mahnke K, and Enk AH. ERK/p38 MAP-kinases and PI3K are involved in the differential regulation of B7-H1 expression in DC subsets. Eur J Immunol. 2010;40(1):254–66.

51. Su S, Zhao J, Xing Y, Zhang X, Liu J, Ouyang Q, et al. Immune Checkpoint Inhibition Overcomes ADCP-Induced Immunosuppression by Macrophages. Cell. 2018;175(2):442–57 e23.

52. Kusmartsev S, Eruslanov E, Kubler H, Tseng T, Sakai Y, Su Z, et al. Oxidative stress regulates expression of VEGFR1 in myeloid cells: link to tumor-induced immune suppression in renal cell carcinoma. J Immunol. 2008;181(1):346–53.

53. Yang L, DeBusk LM, Fukuda K, Fingleton B, Green-Jarvis B, Shyr Y, et al. Expansion of myeloid immune suppressor Gr+CD11b+ cells in tumor-bearing host directly promotes tumor angiogenesis. Cancer Cell. 2004;6(4):409–21.

54. Hickey RJ, Zhou X, Pierson JD, Ravel J, and Forney LJ. Understanding vaginal microbiome complexity from an ecological perspective. Transl Res. 2012;160(4):267–82.

55. Enioutina EY, Bareyan D, and Daynes RA. A role for immature myeloid cells in immune senescence. J Immunol. 2011;186(2):697–707.

56. Salminen A, Kaarniranta K, and Kauppinen A. The role of myeloid-derived suppressor cells (MDSC) in the inflammaging process. Ageing Res Rev. 2018;48:1–10.

57. Dong G, You M, Fan H, Ji J, Ding L, Li P, et al. 17beta-estradiol contributes to the accumulation of myeloid-derived suppressor cells in blood by promoting TNF-alpha secretion. Acta Biochim Biophys Sin (Shanghai). 2015;47(8):620–9.

58. 58. Reed BG, and Carr BR. In: Feingold KR, Anawalt B, Blackman MR, Boyce A, Chrousos G, Corpas E, et al. eds. Endotext. South Dartmouth (MA); 2000.

59. Li P, Chen Y, Xiang Y, Guo R, Li X, Liu J, et al. 17beta-estradiol promotes myeloid-derived suppressor cells functions and alleviates inflammatory bowel disease by activation of Stat3 and NF-kappaB signalings. J Steroid Biochem Mol Biol. 2024;242:106540.

60. Muzny CA, Cerca N, Elnaggar JH, Taylor CM, Sobel JD, and Van Der Pol B. State of the Art for Diagnosis of Bacterial Vaginosis. J Clin Microbiol. 2023;61(8):e0083722.

61. Achkar JM, and Fries BC. Candida infections of the genitourinary tract. Clin Microbiol Rev. 2010;23(2):253–73.

62. Dong M, Wang C, Li H, Yan Y, Ma X, Li H, et al. Aerobic Vaginitis Diagnosis Criteria Combining Gram Stain with Clinical Features: An Establishment and Prospective Validation Study. Diagnostics (Basel). 2022;12(1).

63. Rabasa J, Bradbury M, Sanchez-Iglesias JL, Guerrero D, Forcada C, Alcalde A, et al. Evaluation of the intraoperative human papillomavirus test as a marker of early cure at 12 months after electrosurgical excision procedure in women with cervical high-grade squamous intraepithelial lesion: a prospective cohort study. BJOG. 2020;127(1):99–105.

64. Cantero J, and Genesca M. Maximizing the immunological output of the cervicovaginal explant model. J Immunol Methods. 2018;460:26–35.

65. Scidmore MA. Cultivation and Laboratory Maintenance of Chlamydia trachomatis. Curr Protoc Microbiol. 2005;Chapter 11:Unit 11A 1.

