## Supplementary Materials for "Cervical mucosal inflammation expands functional polymorphonuclear myeloid-derived suppressor cells"

Pieren et al.

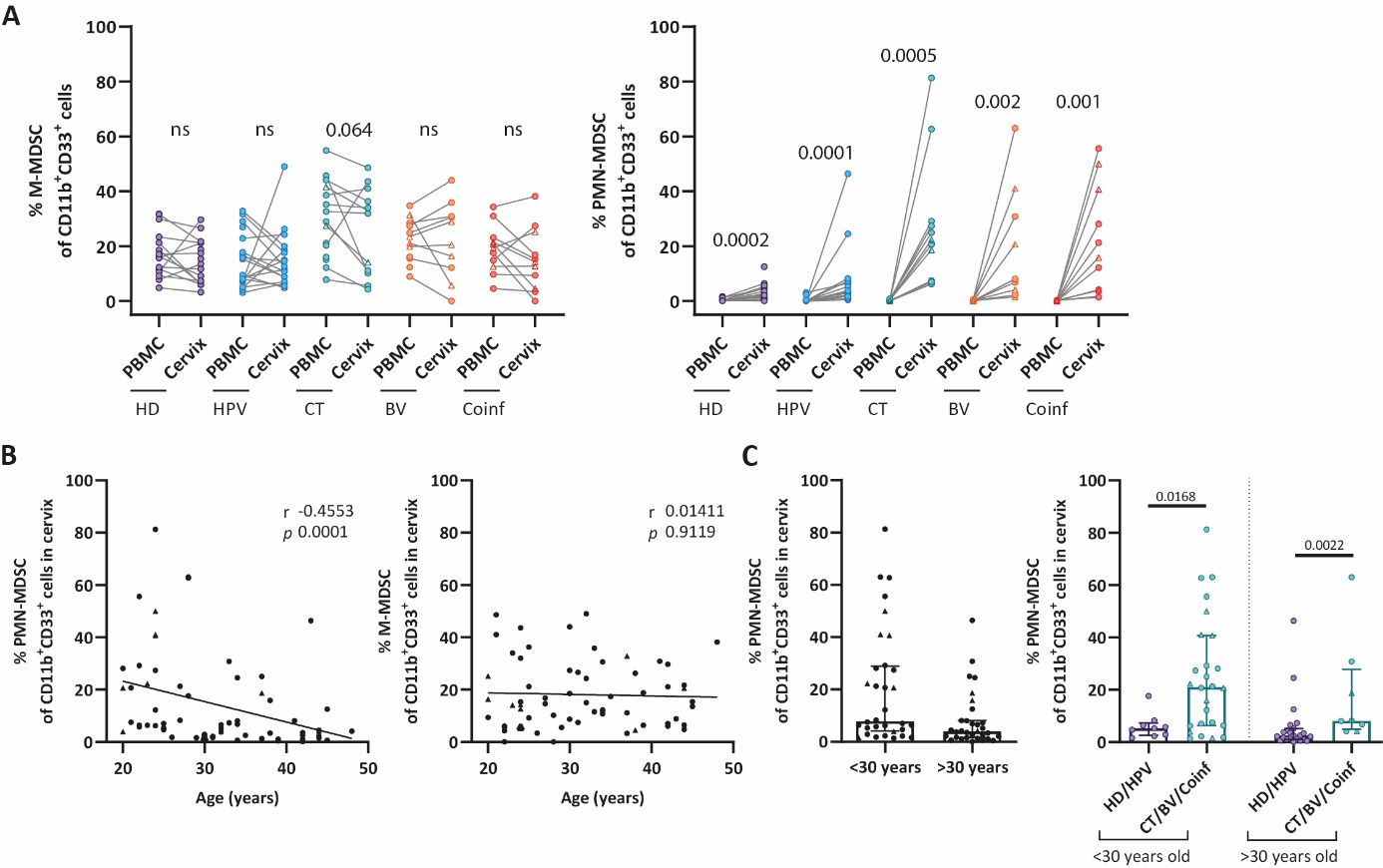

**Supplemental Figure 1. *Frequencies of MDSC subsets in relation to age.***

(A) Comparison of M-MDSC and PMN-MDSC frequencies amongst CD11b^+^CD33^+^ myeloid cells found in paired PBMC and cervical samples within each patient by group (HD n=18; HPV n=15; CT n=14; BV n=12; Coinf n=11). (B) Graphs show the relationship between the frequency of cervical PMN-MDSCs (n=64) and M-MDSCs (n=63) amongst CD11b^+^CD33^+^ myeloid cells with age across all women included in this study. (C) Comparison of cervical PMN-MDSC frequencies amongst CD11b^+^CD33^+^ myeloid cells in women grouped by age (younger n=34 and older n=31 than the median age of 30 years). Data are shown as paired samples (A), correlations (*r* and *P* values) assessed by Spearman test (two-sided) (B), and bar graphs with median and interquartile range (C). Patients with leukocyte/HPF counts of >10 are indicated with triangles. Statistical significance was determined by Wilcoxon test (two-sided) (A), Kruskal-Wallis test with Dunn’s post hoc test (two-sided), or Mann-Whitney test (B, C). Exact *P*-values are shown, ns not significant.

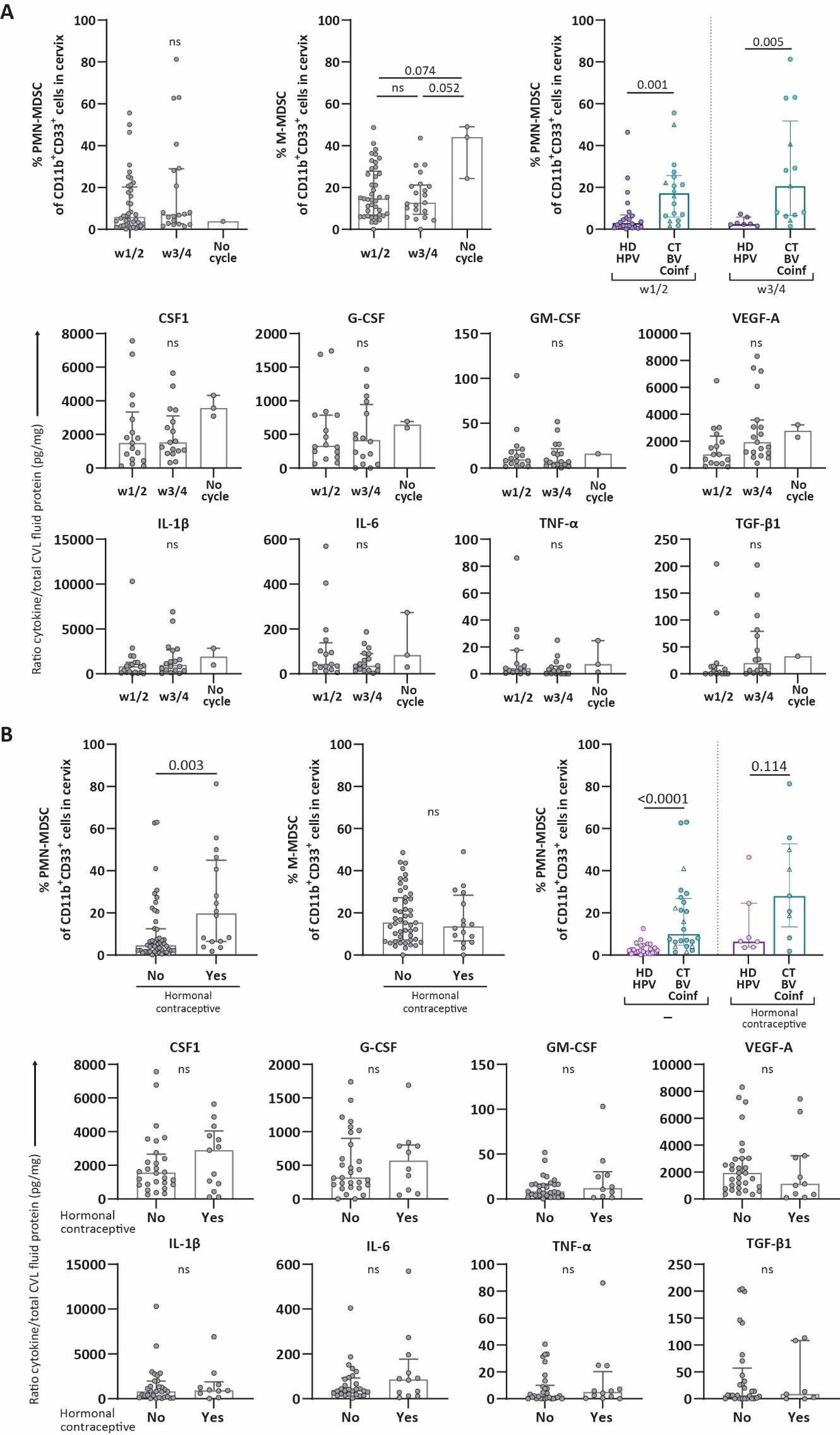

**Supplemental Figure 2. *Frequencies of MDSC subsets and cytokines in CVL fluid in relation to menstrual cycle and hormonal contraception.***

(A) Comparison of cervical PMN-MDSC (week 1/2 n=40; week 3/4 n=20, no cycle n=1) and M-MDSC (week 1/2 n=41; week 3/4 n=21, no cycle n=3) frequencies amongst CD11b^+^CD33^+^ myeloid cells in women grouped by menstrual cycle stage only and grouped by menstrual cycle stage combined with type of infection. Graphs below compare levels of eight different growth factors and cytokines found in CVL fluid of women grouped by menstrual cycle stage, depicted as the ratio of each cytokine per total protein present in the cervical lavage sample (HD n=14; HPV n=12; CT n=11; BV n=11; Coinf n=9). (B) Comparison of cervical PMN-MDSC (no hormonal contraception n=50; with hormonal contraception n=16) and M-MDSC (no hormonal contraception n=48; with hormonal contraception n=16) frequencies amongst CD11b^+^CD33^+^ myeloid cells in women grouped use of hormonal contraception, and grouped by use of hormonal contraception combined with type of infection. Graphs below compare levels of eight different growth factors and cytokines found in CVL fluid of women grouped use of hormonal contraception, depicted as the ratio of each cytokine per total protein present in the cervical lavage sample (HD n=14; HPV n=12; CT n=11; BV n=11; Coinf n=9). Data are shown as bar graphs with median and interquartile range (A). Statistical significance was determined by Kruskal-Wallis test with Dunn’s post hoc test (two-sided) (A), or Mann-Whitney test (B). Exact *P*-values are shown, ns not significant.

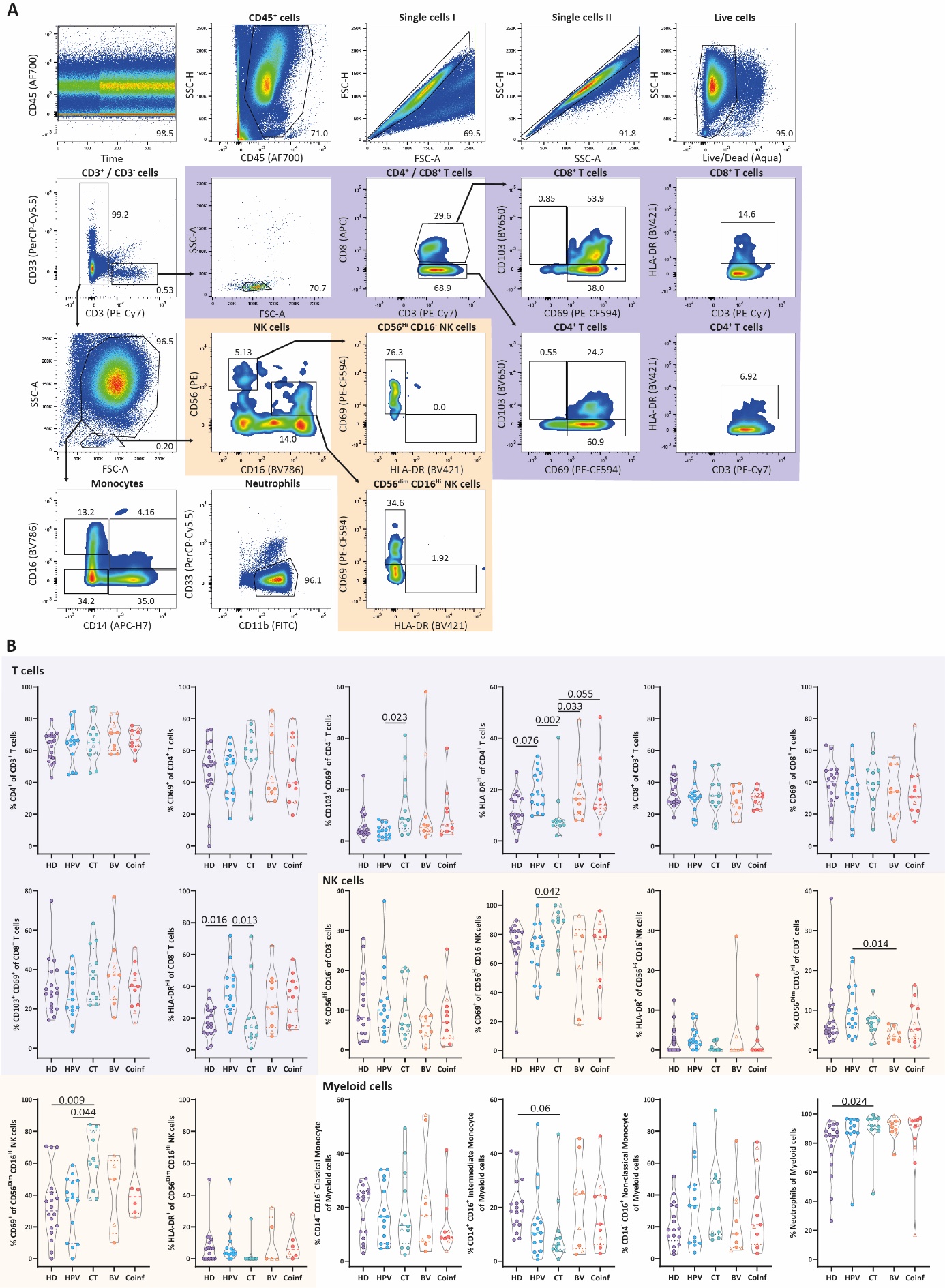

**Supplemental Figure 3. *Cellular subsets in cervix samples from included study participants.***

(A) Representative flow-cytometry plots showing the gating strategy of several cell subsets present in cervical samples. All cellular subsets were identified by gating of time (to exclude disturbances in flow measurements), followed by gating of all CD45^+^, single, live cells (excluding dead and CD20^+^ cells) and subsequent division into CD33^+^ and CD3^+^ cells. Gating on lymphocytes allowed identification of CD4^+^ T cells, CD8^+^ T cells, and NK cells, as well as expression of CD69, CD103, and HLA-DR within these subsets. Gating on myeloid cells allowed identification of classical monocytes, intermediate monocytes, non-classical monocytes, and neutrophils. (B) Comparison of cellular subsets found in cervical samples within each patient group (HD n=18; HPV n=13; CT n=12; BV n=10; Coinf n=11). Data are shown as violin plots with median and quartiles. Patients with leukocyte/HPF counts of >10 are indicated with triangles. Statistical significance was determined by Kruskal-Wallis test with Dunn’s post hoc test (two-sided). *p<0.05; **p<0.01; ***p<0.001; ***p<0.0001.

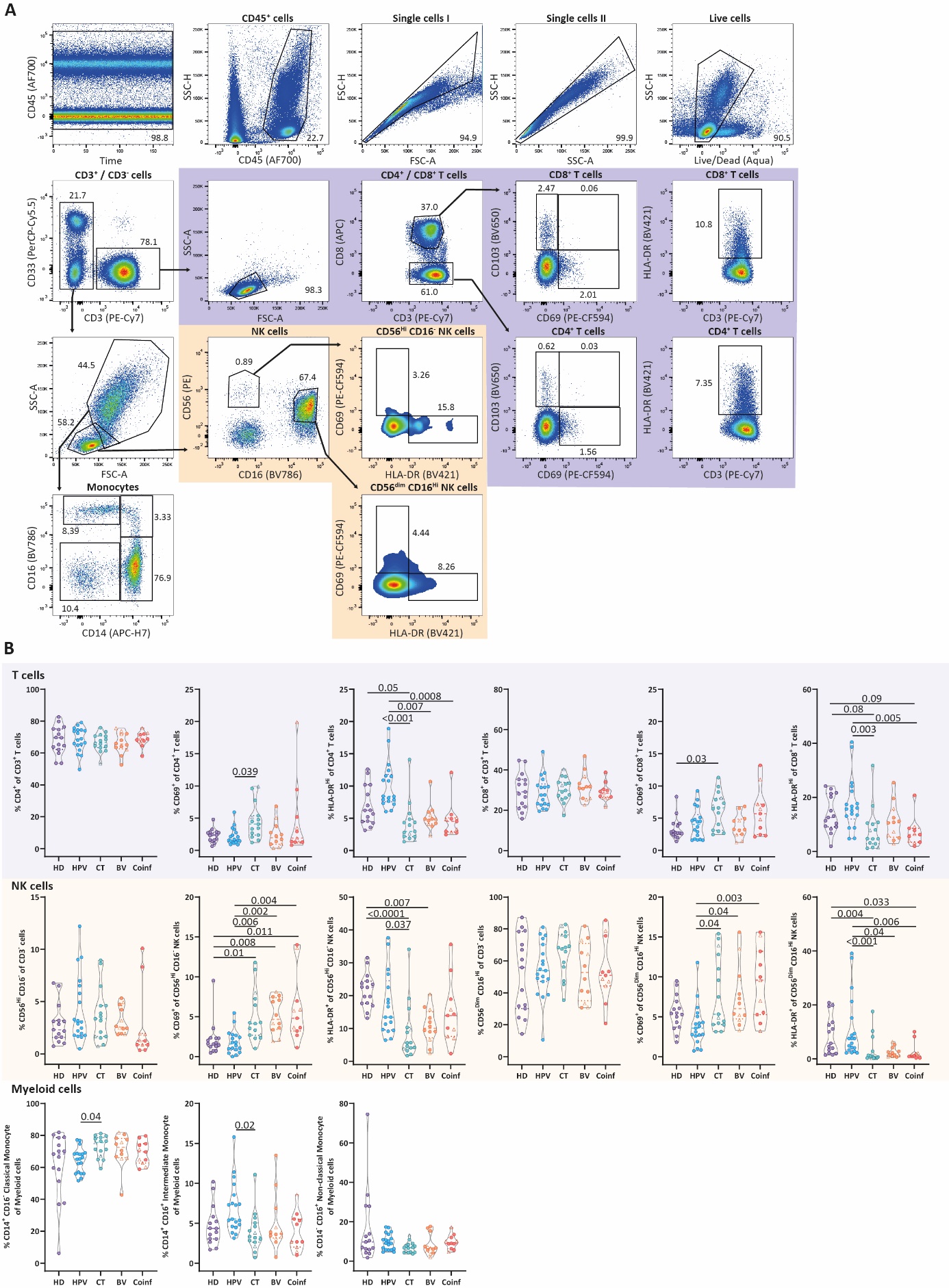

**Supplemental Figure 4. *Cellular subsets in PBMCs samples from included study participants.***

(A) Representative flow-cytometry plots showing the gating strategy of several cell subsets present in PBMC samples. All cellular subsets were identified by gating of time (to exclude disturbances in flow measurements), followed by gating of all CD45^+^, single, live cells (excluding dead and CD20^+^ cells) and subsequent division into CD33^+^ and CD3^+^ cells. Gating on lymphocytes allowed identification of CD4^+^ T cells, CD8^+^ T cells, and NK cells, as well as expression of CD69, CD103, and HLA-DR within these subsets. Gating on myeloid cells allowed identification of classical monocytes, intermediate monocytes, non-classical monocytes. (B) Comparison of cellular subsets found in PBMCs within each patient group (HD n=15; HPV n=20; CT n=14; BV n=12; Coinf n=11). Data are shown as violin plots with median and quartiles. Patients with leukocyte/HPF counts of >10 are indicated with triangles. Statistical significance was determined by Kruskal-Wallis test with Dunn’s post hoc test (two-sided). *p<0.05; **p<0.01; ***p<0.001; ***p<0.0001.

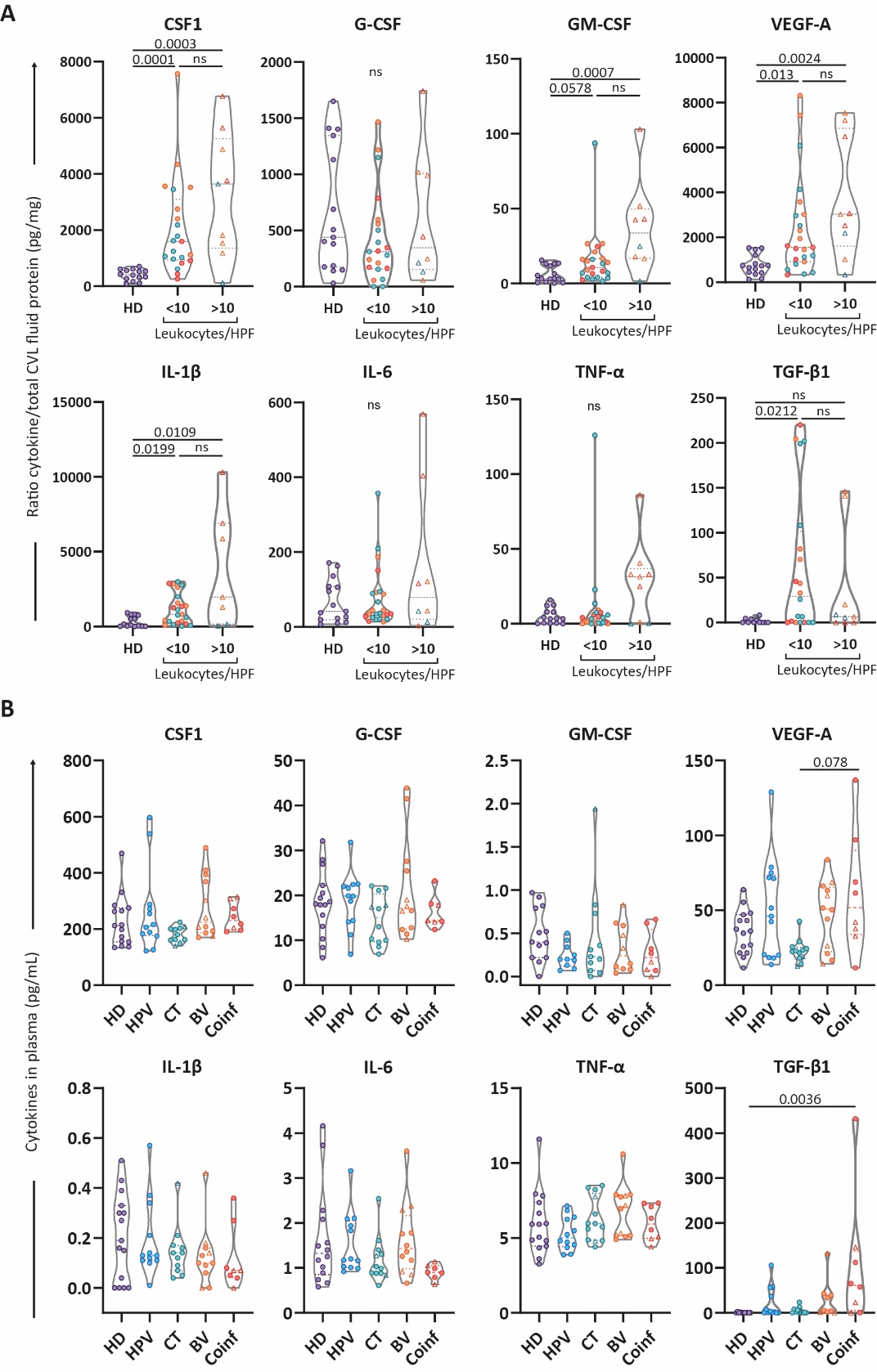

**Supplemental Figure 5. *Cytokine and growth factor levels in plasma show minor differences between groups.***

(A) Comparison of levels of eight different cytokines and growth factors found in CVL fluid (depicted as the ratio of each cytokine per total protein present in the cervical lavage sample) in groups according to leukocyte/HPF count (HD n=18; <10 leukocytes/HPF n=21; >10 leukocytes/HPF n=9). (B) Comparison of levels of eight different cytokines and growth factors found in plasma for each group of women included in this study (HD n=15; HPV n=13; CT n=12; BV n=11; Coinf n=9). Data are shown as violin plots with median and quartiles. Patients with leukocyte/HPF counts of >10 are indicated with triangles. Statistical significance was determined by Kruskal-Wallis test with Dunn’s post hoc test (two-sided). *p<0.05; ***p<0.001.

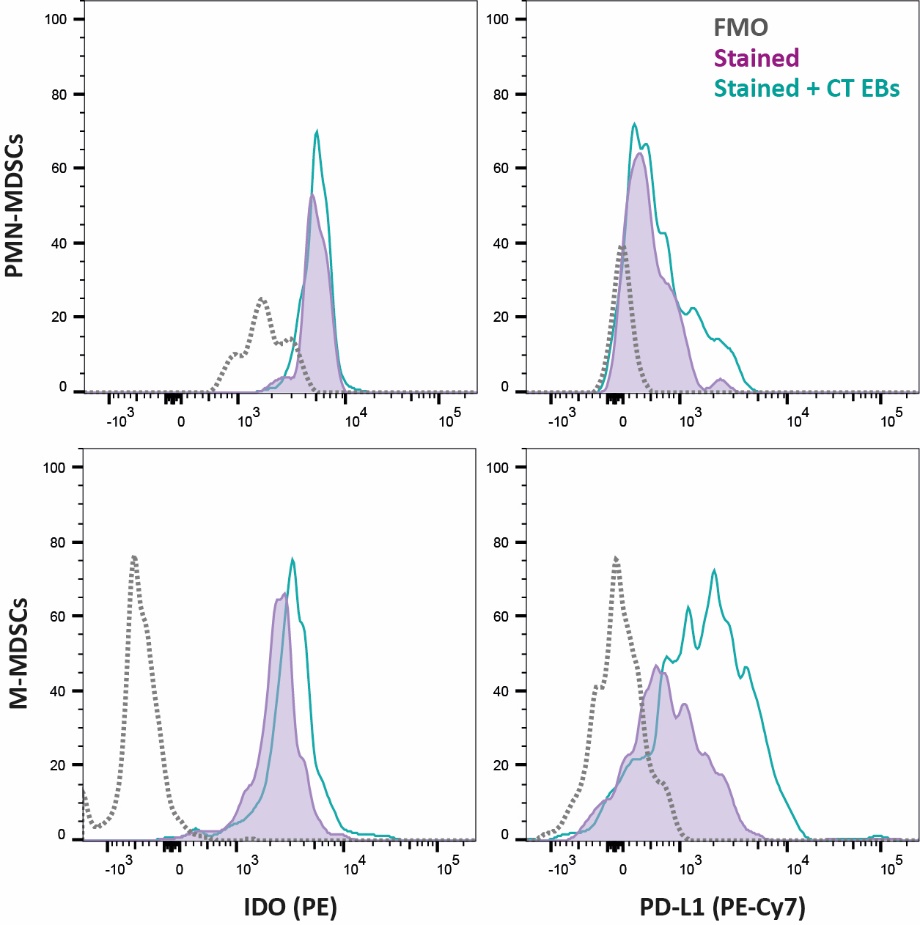

**Supplemental Figure 6. *FMO control for IDO and PD-L1 expression by PMN- and M-MDSCs.***

Histograms show expression of IDO and PD-L1 by PMN-MDSCs and M-MDSCs after staining of CT EB-stimulated cells (Green), unstimulated cells (Purple), and their respective fluorescence minus one (FMO) control (Gray).

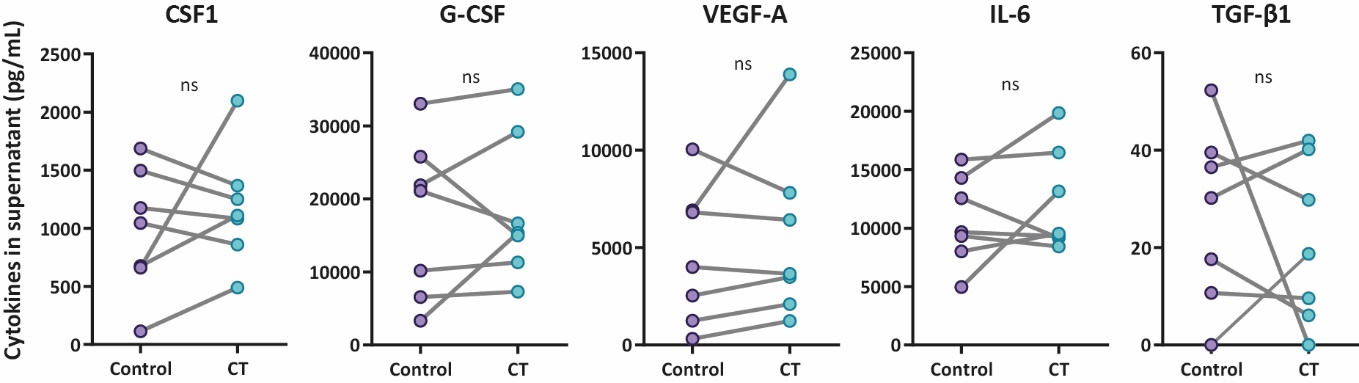

**Supplemental Figure 7. *Cytokine and growth factor levels in supernatant of ex vivo exposed cervical tissue to C. trachomatis elementary bodies.***

Comparison of levels of eight different cytokines and growth factors found in culture supernatant after *ex vivo* exposure of cervical tissue to *C. trachomatis* elementary bodies for 48 hours compared to control (n=7). Data are shown as paired samples. Statistical significance was determined by Wilcoxon test (two-sided).

| **Supplementary Table 1. Characteristics of women with and without genital tract condition included in this study.** | | | | | | | |
| --- | --- | --- | --- | --- | --- | --- | --- |
|  |  | **Healthy** | **HPV** | **CT** | **BV** | **Coinfection** | ***P* value** |
| **Women included in the study (n)** |  | **18** | **19** | **14** | **12** | **11** | **between groups** |
| PBMC and plasma sample acquired, n (%) |  | **15** (83%) | **19** (100%) | **14** (100%) | **12** (100%) | **11** (100%) |  |
| Cytobrush and Lavage sample acquired, n (%) |  | **18** (100%) | **19** (100%) | **13** (93%) | **12** (100%) | **11** (100%) |  |
| **Age (years), median [min.-max.]** |  | **36** [22-45] | **32** [25-44] | **24** [20-37] | **32** [21-44] | **24** [20-48] | **<0.01^a^** |
| **Menstrual cycle, n (%)** |  |  |  |  |  |  | **0.03^b^** |
| S1/S2 |  | 15 (83%) | 10 (53%) | 4 (29%) | 3 (25%) | 7 (64%) |  |
| S3/S4 |  | 3 (17%) | 6 (32%) | 6 (43%) | 7 (58%) | 3 (27%) |  |
| Amenorrhea |  | 0 (0%) | 2 (10%) | 0 (0%) | 1 (8%) | 0 (0%) |  |
| Unknown |  | 0 (0%) | 1 (5%) | 4 (29%) | 1 (8%) | 1 (9%) |  |
| **Hormonal contraceptive** |  |  |  |  |  |  | **0.24^b^** |
| Yes |  | 2 (11%) | 7 (37%) | 2 (14%) | 2 (17%) | 4 (36%) |  |
| No |  | 16 (89%) | 12 (63%) | 12 (86%) | 10 (83%) | 7 (64%) |  |
| **Clinical results, n (%)** |  |  |  |  |  |  |  |
| Prior genital tract infection (<5 months) |  | 0 (0%) | 2 (10%) | 0 (0%) | 1 (7%) | 0 (0%) |  |
| Prior genital tract infection (<1 year) |  | 0 (0%) | 2 (10%) | 1 (7%) | 2 (14%) | 2 (18%) |  |
| HPV genotype: 16, 18/other |  | - | 14/5 (74%/26%) | - | - | - |  |
| Cytomorphologic diagnosis (HSIL/LSIL/ASCUS) |  | - | 12/4/3 (63%/21%/16%) | - | - | - |  |
| Leukocytes/HPF count >10 |  | - | - | 2 (17%) | 4 (33%) | 4 (36%) |  |
| Bacterial vaginosis^#^ |  | - | - | 0 (0%) | 12 (100%) | 10 (91%) |  |
| *Chlamydia trachomatis* |  | - | - | 14 (100%) | 0 (0%) | 8 (73%) |  |
| *Mycoplasma genitalium* |  | - | - | 0 (0%) | 0 (0%) | 1 (9%) |  |
| *Neisseria gonorrhoeae* |  | - | - | 0 (0%) | 0 (0%) | 1 (9%) |  |
| *Trichomonas vaginalis* |  | - | - | 0 (0%) | 0 (0%) | 0 (0%) |  |
| ^#^ Based on ISON-Hay grade 3 |  |  |  |  |  |  |  |
| ^a^ Kruskal-Wallis test with Dunn’s post-test |  |  |  |  |  |  |  |
| ^b^ Chi-square test |  |  |  |  |  |  |  |

| **Supplementary Table 2. Pathogens identified in the coinfection patient group.** | | | |
| --- | --- | --- | --- |
| **Patient** | **Infection #1** | **Infection #2** | **Infection #3** |
| 1 | *Chlamydia trachomatis* | Bacterial Vaginosis | - |
| 2 | *Chlamydia trachomatis* | Bacterial Vaginosis | - |
| 3 | *Chlamydia trachomatis* | Bacterial Vaginosis | - |
| 4 | *Chlamydia trachomatis* | Bacterial Vaginosis | - |
| 5 | *Chlamydia trachomatis* | Bacterial Vaginosis | - |
| 6 | *Chlamydia trachomatis* | Bacterial Vaginosis | - |
| 7 | *Chlamydia trachomatis* | *Candida albicans* | - |
| 8 | *Chlamydia trachomatis* | Bacterial Vaginosis | *Candida albicans* |
| 9 | Bacterial Vaginosis | *Candida albicans* | - |
| 10 | Bacterial Vaginosis | *Candida albicans* | - |
| 11 | Bacterial Vaginosis | *Neisseria gonorrhoeae* | *Mycoplasma genitalium* |

**Supplementary Table 3. *Correlations between the frequency of cervical PMN-MDSCs or M-MDSCs and cytokine and growth factor levels in CVL fluid***

|  | **% PMN-MDSC of CD11b+CD33+ cells** | **G-CSF** | **CSF1** | **IL-1β** | **GM-CSF** | **IL-6** | **VEGF-A** | **TNF-α** | **TGF-β1** |
| --- | --- | --- | --- | --- | --- | --- | --- | --- | --- |
| **Total** | r | -0.198 | **0.459** | 0.116 | 0.236 | -0.098 | 0.192 | -0.225 | 0.225 |
|  | P (two-tailed) | ns | **0.0009** | ns | 0.1061 | ns | ns | ns | ns |
| **HD** | r | -0.143 | -0.247 | -0.390 | **-0.662** | **-0.881** | -0.222 | **-0.570** | 0.097 |
|  | P (two-tailed) | ns | ns | ns | **0.0117** | **<0.0001** | ns | **0.0452** | ns |
| **HPV** | r | 0.381 | **0.738** | 0.393 | 0.486 | 0.048 | -0.286 | -0.095 | 0.657 |
|  | P (two-tailed) | ns | **0.0458** | ns | ns | ns | ns | ns | ns |
| **CT** | r | 0.236 | 0.249 | **0.709** | 0.420 | 0.346 | 0.109 | 0.334 | 0.174 |
|  | P (two-tailed) | ns | ns | **0.0182** | ns | ns | ns | ns | ns |
| **BV** | r | -0.183 | 0.417 | -0.467 | 0.500 | 0.333 | **0.733** | -0.667 | 0.381 |
|  | P (two-tailed) | ns | ns | ns | ns | ns | **0.0202** | ns | ns |
| **Coinf** | r | -0.167 | -0.083 | -0.214 | 0.200 | 0.286 | 0.117 | 0.317 | -0.612 |
|  | P (two-tailed) | ns | ns | ns | ns | ns | ns | ns | ns |
|  | **% M-MDSC of CD11b+CD33+ cells** | **G-CSF** | **CSF1** | **IL-1β** | **GM-CSF** | **IL-6** | **VEGF-A** | **TNF-α** | **TGF-β1** |
| **Total** | r | -0.081 | 0.147 | -0.110 | -0.063 | -0.041 | -0.049 | -0.080 | -0.102 |
|  | P (two-tailed) | ns | ns | ns | ns | ns | ns | ns | ns |
| **HD** | r | -0.139 | **-0.742** | -0.352 | -0.172 | -0.081 | -0.196 | -0.124 | -0.321 |
|  | P (two-tailed) | ns | **0.0051** | ns | ns | ns | ns | ns | ns |
| **HPV** | r | -0.107 | **0.881** | 0.500 | 0.371 | 0.333 | 0.107 | 0.548 | **1.000** |
|  | P (two-tailed) | ns | **0.0072** | ns | ns | ns | ns | ns | **0.0167** |
| **CT** | r | -0.115 | -0.358 | -0.591 | -0.292 | -0.188 | -0.191 | -0.237 | -0.468 |
|  | P (two-tailed) | ns | ns | 0.0609 | ns | ns | ns | ns | ns |
| **BV** | r | 0.417 | 0.300 | 0.183 | -0.133 | -0.033 | -0.479 | 0.037 | -0.548 |
|  | P (two-tailed) | ns | ns | ns | ns | ns | ns | ns | ns |
| **Coinf** | r | -0.286 | -0.033 | -0.286 | -0.417 | 0.238 | -0.150 | -0.183 | 0.408 |
|  | P (two-tailed) | ns | ns | ns | ns | ns | ns | ns | ns |

Spearman rank correlation
